# Evaluating Clinical Foundation Models for Early Alzheimer’s Disease and Related Dementia Prediction from Longitudinal EHRs

**DOI:** 10.64898/2026.09.01.26361933

**Authors:** Shahla Farzana, Ash Arian, Tatjana Rundek, Moïse Desvarieux, Habibul Ahsan

**Affiliations:** Institute for Population and Precision Health, University of Chicago, Chicago, IL, USA; Pritzker School of Medicine, University of Chicago, Chicago, IL, USA; Department of Neurology, Miller School of Medicine, University of Miami, Miami, Florida, USA; Department of Epidemiology, Mailman School of Public Health, Columbia University, New York, USA; Department of Public Health Sciences, University of Chicago, Chicago, IL, USA; Department of Family Medicine, Biological Sciences Division, University of Chicago Medicine, Chicago, IL, USA

## Abstract

Early identification of Alzheimer’s disease and related dementias (ADRD) remains challenging despite its importance for timely intervention, management of modifiable risk factors, and care planning. We developed and evaluated ADRD onset prediction models using longitudinal electronic health records (EHRs) from the All of Us Research Program at clinically meaningful lead times of 6, 12, 24, and 36 months before diagnosis, benchmarking interpretable count-based representations against four publicly available pretrained clinical foundation models (CLMBR-T, GPT-style, LLaMA-style, and Mamba) across multiple ADRD phenotype definitions. Count-based models consistently achieved the highest discrimination and calibration across all cohorts and prediction horizons. Predictive performance declined with increasing lead time for all approaches; however, the performance gap between count-based and pretrained representations progressively narrowed, with foundation models achieving comparable AUROC of 0.719 (compared to the AUROC of 0.738 of count-based model) at the 36-month horizon while providing higher sensitivity and F1 scores under a fixed operating threshold. External validation with zero-shot evaluation on UChicago EHRs exhibited limited generalizability for count-based and pretrained clinical foundation model based representations. These findings demonstrate that transparent count-based EHR representations remain the strongest overall approach for ADRD onset prediction, while pretrained clinical foundation models provide complementary advantages for long-term risk identification and establish a benchmark for evaluating transferable clinical representations in temporal ADRD risk prediction.

## 1 Introduction

Alzheimer’s disease and related dementias (ADRD) affect more than six million people 65 years and older in the United States and remain among the leading causes of disability and mortality in older adults [1]. Because neuropathological changes begin years before a clinical diagnosis, identifying individuals at elevated risk during the prodromal phase offers opportunities for earlier intervention, management of modifiable cardiovascular and lifestyle risk factors, care planning, recruitment into clinical trials, and initiation of emerging disease-modifying therapies. However, early recognition in routine clinical practice remains challenging because cognitive symptoms are often subtle, heterogeneous, and under-recognized, particularly among underserved populations.

Electronic health records (EHRs) provide an attractive platform for scalable ADRD risk assessment because they capture longitudinal healthcare utilization, diagnoses, medications, laboratory measurements, procedures, and demographic information generated during routine clinical care. Unlike neuroimaging, blood biomarkers, or dedicated cognitive assessments, EHR-based prediction models can be deployed passively within existing healthcare systems without requiring additional testing or patient burden. Consequently, numerous machine learning approaches have been developed for ADRD prediction using structured EHR data. Early work demonstrated that tree-based machine learning models substantially outperform traditional knowledge-driven risk models by leveraging high-dimensional longitudinal EHR features [2]. More recent studies have further incorporated temporal modeling, knowledge networks, interpretable dynamic risk prediction, and foundation-model representations to improve predictive performance and characterize evolving risk over time [3, 4, 5].

Recent advances in clinical foundation models have introduced pretrained representations of longitudinal EHR trajectories that can be transferred to downstream prediction tasks with minimal task-specific training. Large multimodal foundation models such as RisQ [6] learn shared disease representations across multiple diseases, modalities, and time using large biobank cohorts, whereas recent large language model (LLM)-based approaches have demonstrated improved ADRD prediction by augmenting conventional supervised learning pipelines with general-purpose language models [7]. Although these approaches demonstrate the promise of representation learning, it remains unclear whether pretrained clinical representations consistently outperform transparent count-based EHR models for a single clinically important endpoint such as ADRD onset, particularly under standardized OMOP-based pipelines and across multiple clinically meaningful lead times. Several important gaps therefore remain. First, most studies evaluate prediction at a single horizon or optimize continuously updated dynamic risk estimates, making it difficult to understand how predictive performance evolves as diagnosis approaches. Second, relatively few studies directly compare interpretable count-based EHR representations with off-the-shelf pretrained clinical foundation models using identical patient cohorts and evaluation protocols. Third, despite increasing interest in model interpretability, comparatively little is known about how reliance on specific clinical features changes from early to late stages preceding ADRD diagnosis. Finally, external validation across independent healthcare systems remains limited for many recently proposed approaches.

In this study, we evaluate whether routinely collected longitudinal EHR data can identify patients who subsequently develop ADRD at four clinically meaningful prediction horizonsâ36, 24, 12, and 6 months before diagnosisâand characterize how predictive performance and clinically relevant predictors evolve across these lead times. Our objective complements recent dynamic prediction frameworks that continuously update patient risk as new clinical information becomes available by instead evaluating fixed lead-time prediction windows that facilitate systematic comparison of prediction difficulty and feature importance across disease progression.

Specifically, this work makes four primary contributions:

- **Generalizability.** We evaluate the proposed framework across two independent health systems using standardized OMOP Common Data Model pipelines and multiple ADRD phenotype definitions.
- **Temporal characterization.** We systematically compare predictive performance, calibration, and clinically interpretable feature importance across four clinically meaningful prediction horizons (36, 24, 12, and 6 months before diagnosis).
- **Representation learning benchmark.** We provide one of the first comprehensive evaluations of publicly available pretrained clinical foundation model representations (CLMBR, GPT-style, Llama-style, and Mamba encoders) against transparent count-based EHR models for ADRD onset prediction under identical cohorts, prediction horizons, and evaluation protocols.
- **Integration of survey-derived risk factors.** We systematically evaluate whether temporally aligned self-reported lifestyle and personal/family health history variables from the All of Us Research Program provide incremental predictive value beyond routinely collected longitudinal EHR data, quantifying their contribution across model families and prediction horizons.

**Figure 1:**
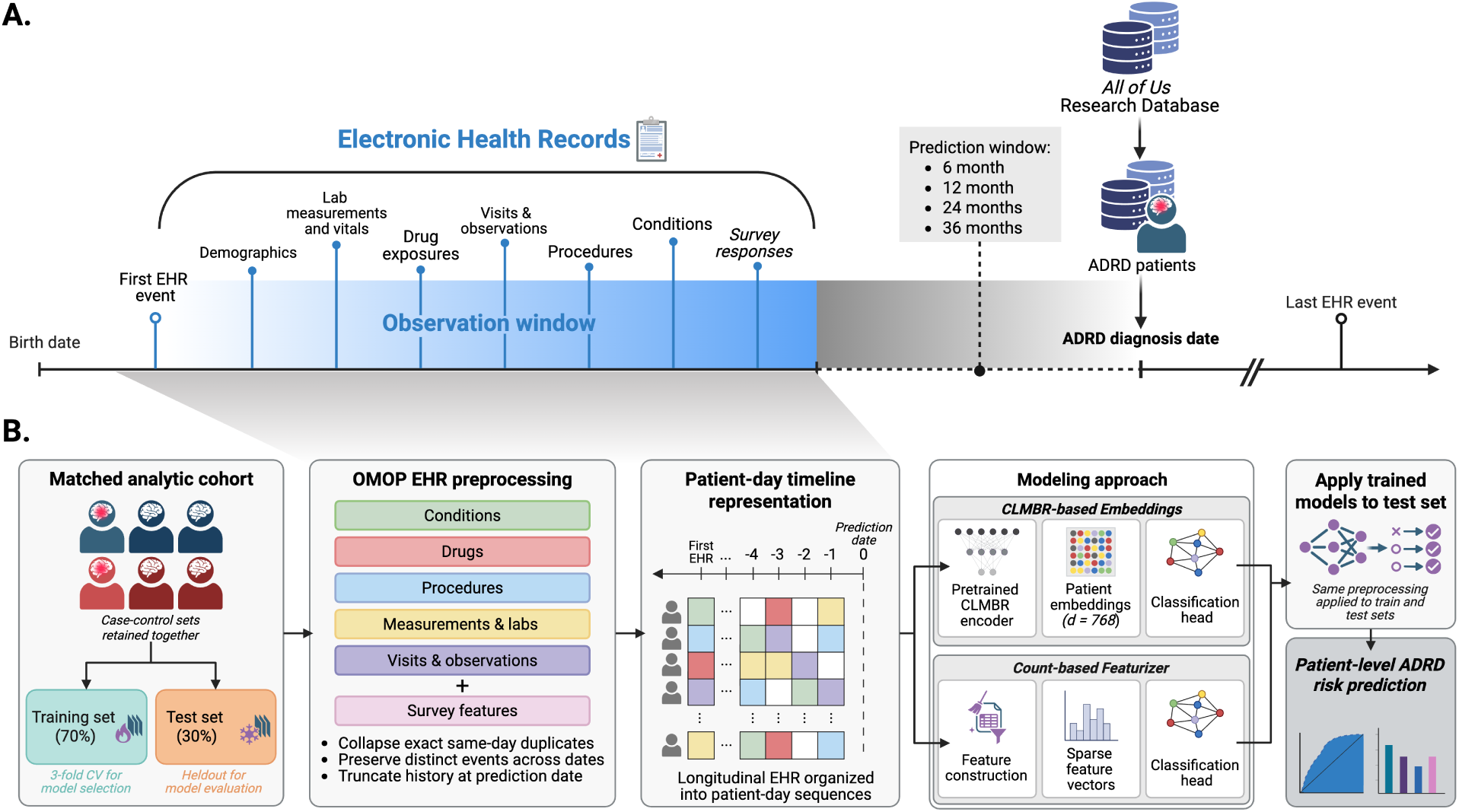
Study design and modeling framework for EHR-based ADRD onset prediction. **(A)** Temporal design of the prediction task, defining the longitudinal observation period and prediction date relative to ADRD onset; only information recorded before the prediction date was available to the models. **(B)** End-to-end modeling workflow in which matched case–control sets were transformed from longitudinal OMOP-format EHR data into patient-day representations and partitioned at the matched-set level to prevent data leakage. Models were trained using either pretrained Clinical Language Model-Based Representations (CLMBR) with a supervised classification head or engineered count-based features with classical machine-learning classifiers. Model and hyperparameter selection were performed exclusively within the training partition, after which the learned mapping between patient representations and ADRD outcome was applied without retraining to the held-out test set to generate patient-level risk estimates.

## 2 Results

### 2.1 Study Cohort Characteristics

Because our CD1 (i.e., have at least one encounter with a valid ADRD diagnosis code or at least one encounter of anti-dementia medication) has the most relaxed rules, Table 1 shows the descriptive statistics of the case and control groups identified using CD1 for All of US (AoU) and UChicago (External validation) datasets. Descriptive statistics of the secondary-confirmed cohort (have at least one encounter with a valid ADRD diagnosis code followed by anti-dementia medication prescription) CD2 are detailed in the Supplementary Table S3. The demographic composition between AoU and UChicago dataset differs substantially. UChicago includes an older population (median index age approximately 79 years vs. 71 years in AoU), a substantially higher proportion of Black patients (62% vs. 13%), fewer White patients (33% vs. 65%), and fewer Hispanic participants. There is also striking difference in clinical history density between two datasets. Compared with AoU, UChicago patients have substantially shorter longitudinal follow-up (median history 8.2 vs. 13.8 years for cases; 7.7 vs. 10.7 years for controls) and markedly fewer recorded encounters (62.4 vs. 325.3 encounters for cases; 55.4 vs. 180.2 for controls). Similar reductions are observed across ambulatory, inpatient, and emergency encounters. These differences indicate that the external validation dataset represents a distinct patient population rather than a simple replication of AoU, providing a rigorous assessment of generalizability.

**Table 1:**
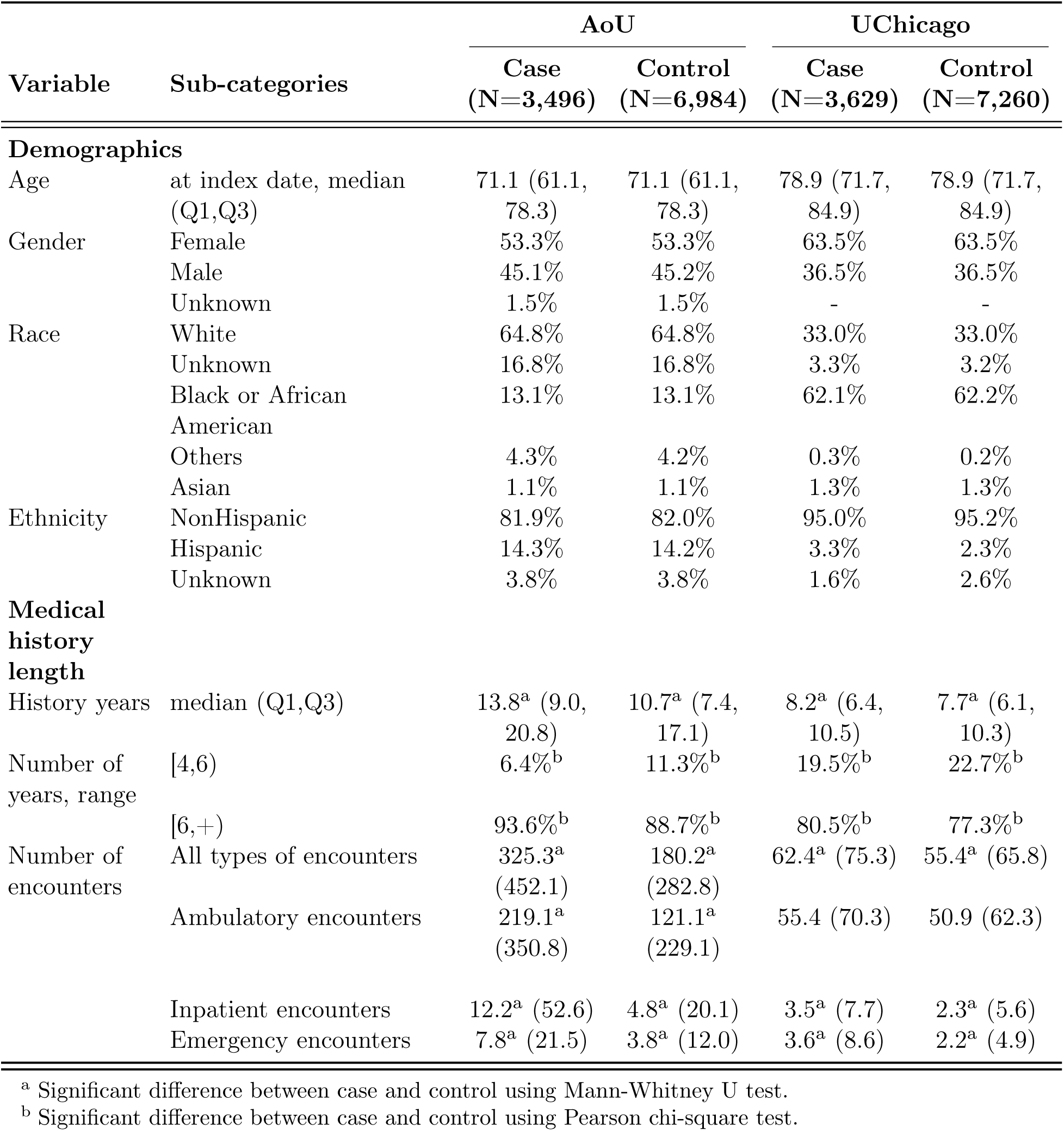
Demographic characteristics, healthcare utilization, and EHR history for CD1 cohort. Number of encounters are represented as *mean (SD)*.

### 2.2 ADRD Risk Prediction Performance

In this subsection, we present the overall results of the experimental settings for ADRD risk prediction.

#### Discriminative performance across prediction horizons

Figure 2 shows the comparative performance of each model family on held-out test set selected by their performance on 3-fold cross validation. Across the four prediction horizons, the count-based model consistently achieved the best overall predictive performance for the CD1 case definition. At the 6-month horizon, it substantially outperformed all clinical foundation models, achieving an AUROC of 0.827 and an AUPRC of 0.722, compared with AUROCs of 0.752â0.784 and AUPRCs of 0.608â0.662 for the embedding-based models. This advantage persisted at the 12-, 24-, and 36-month horizons, although the performance gap gradually narrowed as the prediction horizon increased. By 36 months, the count-based model retained the highest AUROC (0.738 vs. 0.682â0.719) and AUPRC (0.589 vs. 0.517â0.560), but the differences relative to the strongest embedding-based models were notably smaller than at the 6-month horizon. In the secondary-confirmed cohort (CD2), which contained the fewest case and control participants among the three case definitions, all model families exhibited lower discriminative performance than in CD1 across prediction horizons. Unlike the other cohort definitions, discrimination did not decline monotonically with increasing prediction horizon. The highest AUROC and AUPRC were observed at the 12-month horizon for the count-based model (AUROC 0.802, AUPRC 0.690), followed by a gradual decline at the 24-and 36-month horizons. Among embedding-based models, GPT-2048 and LLaMA-512 generally provided the strongest discrimination, with AUROC for GPT-2048 and LLaMA-512 ranging from of 0.767 and 0.759 at 6-month horizon to 0.659 and 0.647 at 36-month horizon respectively. Similar to CD1, the performance gap between count-based and embedding-based approaches narrowed modestly at longer prediction horizons in CD2. External zero-shot validation on the UChicago dataset resulted in lower performance (Supplementary Figure S3) for all model families compared with models trained on UChicago EHR data (Supplementary Figure S2). This performance decline is expected given the substantial differences between two datasets, including older patients, a markedly different racial composition, shorter longitudinal clinical histories, and differences in healthcare utilization patterns in UChicago dataset compared with AoU (Table 1, Supplementary Table S3). In UChicago dataset, the count-based models experienced the largest reduction (compared to the in-domain trained models) in discrimination across prediction horizons. In contrast, the pretrained CLMBR models exhibited comparatively smaller degradation in some settings despite showing more variable performance across prediction horizons, particularly in CD2. Overall, these findings suggest that manually engineered count-based features capture short-term disease signals particularly effectively, while the relative discriminative performance of pretrained clinical foundation models improves for longer-term risk prediction. Across all model families, incorporating survey-derived features produced only marginal improvements over EHR-only models. Across case definitions and prediction horizons, AUROC gains did not exceed 1.4% (for CD1), with similarly modest changes in AUPRC and F1 score. The limited benefit likely reflects the substantial missingness of pre-index survey data, although smoking-related variables contributed the greatest incremental value because they were the most frequently available survey features.

**Figure 2:**
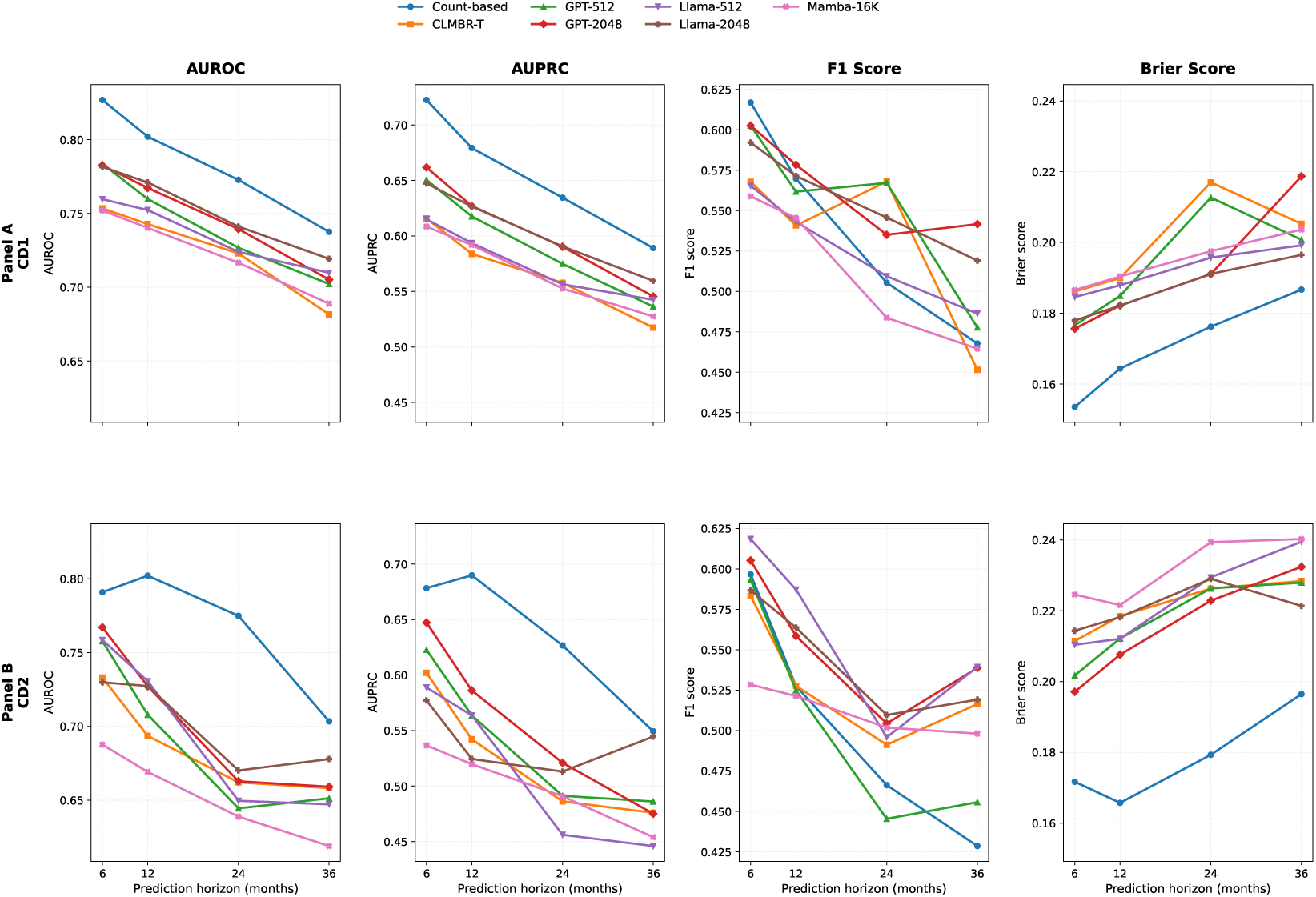
Held-out test set performance of the best-performing feature variant within each model family across all case definitions and prediction horizons for AoU dataset.

#### Threshold-dependent classification performance

In contrast to AUROC and AUPRC, the F1-score showed a different trend across all case definitions. Although the count-based model achieved the highest F1-score at the 6-month horizon (0.617) for CD1, embedding-based models became increasingly competitive at longer prediction horizons. At 24 and 36 months, several foundation models, particularly GPT-2048, and Llama-2048, achieved F1-scores which exceeded those of the count-based model. F1-score exhibited the similar pattern across CD2. For example, while the count-based model achieved the highest F1 score at the 6-month horizon (0.597), its performance declined substantially to 0.429 at 36 months for CD2. In contrast, several embedding-based models maintained relatively stable F1 scores across prediction horizons, with GPT-2048 and LLaMA-512 achieving 0.539 at 36 months, exceeding the count-based model despite lower discriminative performance. Similar trend holds for zero-shot transfer of count-based and embedding based models on UChicago test set (Supplementary Figure S3). These findings suggest that, despite lower overall discriminative performance, foundation-model embeddings provide a more favorable balance between precision and recall under a fixed decision threshold for longer-term prediction. To further evaluate the robustness of model performance across decision thresholds, threshold sensitivity analyses are presented in the Supplementary Figure (S4).

#### Model calibration

Calibration trends were similar across all three case definitions. Models were generally well calibrated at shorter prediction horizons but exhibited progressively poorer calibration as the prediction horizon increased from 6 to 36 months. Correspondingly, Brier scores increased with prediction horizon for all model families. For example, for CD1 at 6 months the brier score for the count-based model vs. the best-performing embedding-based model was 0.169 and 0.189, whereas at 36 months horizon the score increased to 0.198 and 0.225 respectively, indicating greater uncertainty and reduced calibration accuracy for long-term ADRD risk prediction. Similarly, for CD2, The count-based model consistently demonstrated the lowest Brier scores, increasing from 0.172 at 6 months to 0.196 at 36 months, whereas embedding-based models exhibited higher Brier scores throughout (approximately 0.211â0.240). This trend indicates increasing predictive uncertainty for longer-term ADRD risk prediction, while also confirming that count-based models remained better calibrated than embedding-based approaches across all prediction horizons. Reliability diagram for representative models for CD1 are provided in Supplementary Figure (S5), demonstrating generally good agreement between predicted probabilities and observed event rates at shorter prediction horizons, with progressively larger deviations as the prediction horizon increases.

#### Effect of foundation model context length on ADRD prediction performance

Figure 2 compares GPT-512, GPT-2048, Llama-512, and Llama-2048, which differ only in their pretrained maximum context lengths. All models were constructed from the same pre-cutoff clinical histories. For patients whose tokenized histories exceeded the model’s context length, only the most recent events up to the model’s maximum input length (512 or 2048 tokens) were retained. Consequently, this comparison evaluates the combined effect of pretrained architecture and available historical context. Across cohort definitions and prediction horizons, increasing the maximum context length resulted in only modest and architecture-dependent differences. In CD1, Llama-2048 generally achieved slightly higher discriminative performance than Llama-512 across prediction horizons. In CD2, performance differences between the two models were smaller and less consistent; however, Llama-2048 tended to outperform Llama-512 at the longer prediction horizons (24 and 36 months), whereas little difference was observed at the shorter horizons. Similarly, GPT-2048 exhibited only marginal and horizon-dependent differences relative to GPT-512, without a consistent performance advantage across either cohort. These patterns were consistent between cross-validation and held-out test evaluations. Overall, increasing the maximum context length of pretrained models did not consistently improve ADRD risk prediction.

### 2.3 Model Reliance on Clinically Interpretable EHR Features

Figure 3 summarizes the relative contribution of clinically interpretable features (top 10) to the XGBoost models across cohort definitions and prediction horizons, while Supplementary Figure S6 presents the complete set of the top 20 predictors. Across cohorts, memory-related diagnoses (e.g., amnesia and minimal cognitive impairment) became increasingly important closer to the prediction date. Across all cohort definitions, memory-related diagnoses, including amnesia and minimal cognitive impairment, became progressively more influential as the prediction date approached, consistent with increasing manifestation of cognitive symptoms before ADRD diagnosis. In contrast, broader indicators of healthcare utilization, including drug utilization, visit frequency, observation frequency, and diagnosis frequency, contributed relatively more at longer prediction horizons, suggesting that patterns of healthcare engagement capture early risk before disease-specific manifestations emerge. Psychiatric conditions, particularly depressive disorder and mental disorder, showed persistent but modest contributions across horizons, whereas laboratory measurements (e.g., vitamin B12), socioeconomic status (household income), and lifestyle-related variables (smoking history) exhibited smaller and more cohort-specific effects. Additional clinically relevant predictors, including, Parkinson’s disease, dermatological condition appeared among the top 20 predictors for specific cohort definitions or prediction horizons. Overall, these findings suggest that the models assign greater importance to nonspecific indicators of health complexity and healthcare utilization at longer prediction horizons, whereas disease-specific cognitive phenotypes increasingly dominate prediction closer to ADRD onset.

**Figure 3:**
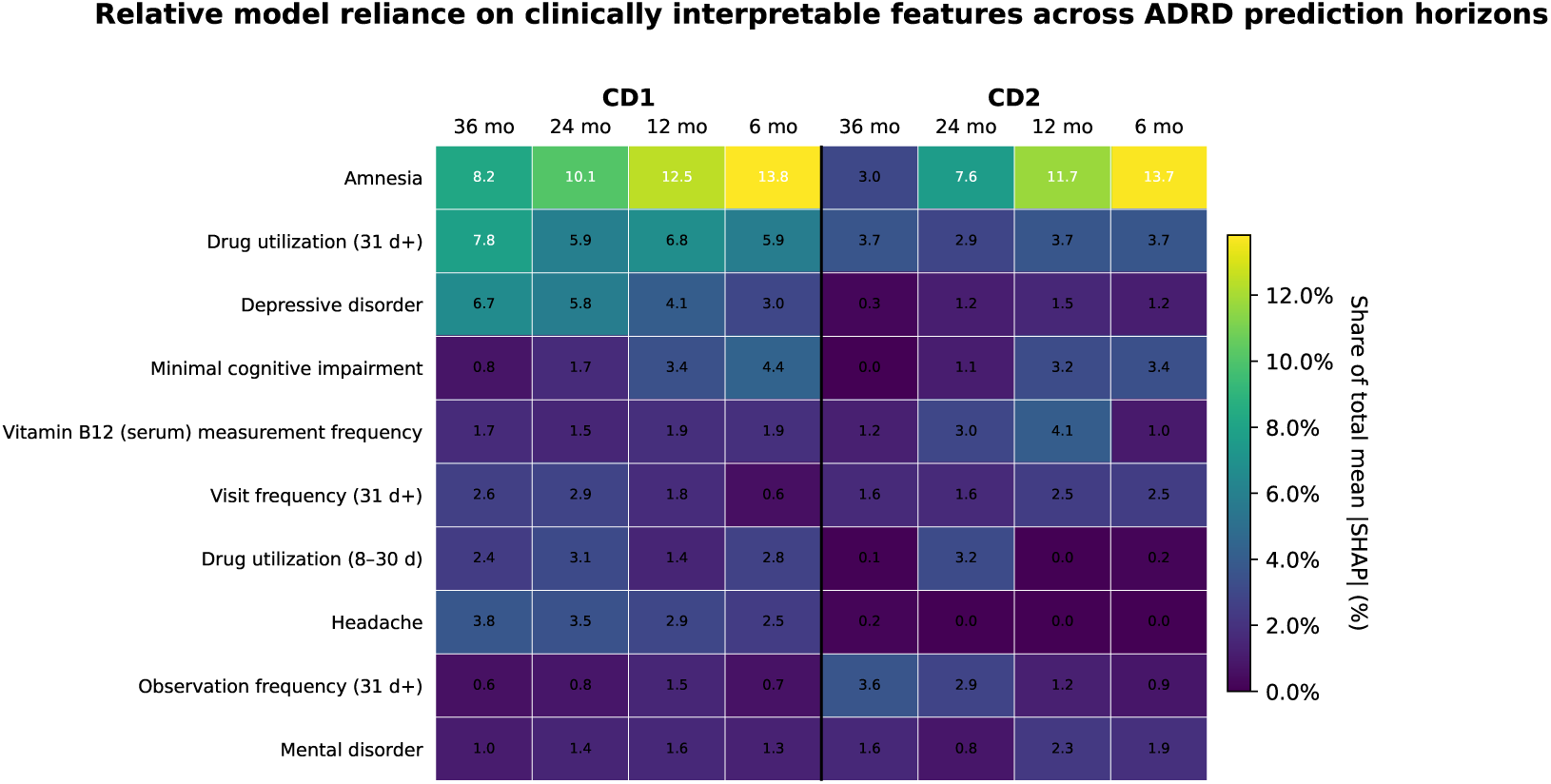
Cross-horizon SHAP heatmap across case definitions for AoU dataset. Cell values represent each feature’s percentage contribution to the total mean absolute SHAP attribution within the corresponding model. Features were selected based on their rankings across case definitions and prediction horizons; however, heatmap values were obtained from the complete feature attribution set for each model, including instances in which a displayed feature did not individually rank among that model’s top 10 predictors. 31 d+ represents all events recorded more than 30 days before the prediction date, whereas 1â7 d and 8â30 d capture more proximal, non-overlapping time windows. A comprehensive heatmap including the top 20 features is provided in Supplementary Figure S6.

#### What do utilization features represent clinically?

Because healthcare utilization features consistently ranked among the most important predictors in the global SHAP analysis (Figure 3), we performed a detailed clinical decomposition of these features in the CD2 (secondary-confirmed) cohort at the 36-month prediction horizon. For the visit-frequency SHAP feature, patient-prevalence analysis indicated that positive model attribution was primarily associated with hospital-, inpatient-, and emergency-related encounters (Supplementary Figure S7 (a)), suggesting that the model captured patterns of intensive healthcare utilization. For the drug-utilization SHAP feature, an initial patient-prevalence analysis showed that nearly all therapeutic classes were less prevalent among patients with high positive SHAP attribution. Examination of the SHAP dependence plot revealed that positive SHAP values were primarily associated with lower overall drug utilization, whereas extensive medication use contributed negatively to the model prediction. Thus, differences in therapeutic-class prevalence largely reflected differences in the overall volume of medication exposure, rather than differences in medication composition. To account for this, we normalized each patient’s medication profile by their total number of drug events and compared the mean within-patient proportion of each therapeutic class between SHAP groups. After normalization, patients with high positive SHAP attribution exhibited a smaller average share of gastrointestinal medications, antidepressants, vitamins/supplements, and antidiabetic medications than patients with low-SHAP patients, whereas differences in the remaining therapeutic classes were not statistically significant after false-discovery-rate correction (Supplementary Figure S7 (b)). These findings suggest that the predictive contribution of the drug-utilization feature was driven primarily by overall medication utilization, with only modest differences in medication-class composition after accounting for total drug exposure.

### 2.4 Performance Across Clinically Relevant Subcohorts

Performance across clinically relevant subgroups was evaluated for the primary CD1 cohort at 6-month prediction horizon (Figure 4), with analyses for the longest prediction horizon (36-month) provided in Supplementary Figure S8. Across all demographic and clinical history subgroups, predictive performance declined as the prediction horizon increased from 6 to 36 months. The count-based model consistently achieved the highest AUROC and AUPRC, followed by the GPT-2048 and Llama-2048 models. Performance was relatively similar between males and females, whereas substantial heterogeneity was observed across age groups, with individuals aged *≥* 85 years exhibiting the lowest discrimination and greatest variability. Racial subgroup differences were modest, with White participants generally demonstrating slightly higher performance than Black and Other participants. Stratification by EHR history revealed a horizon-dependent pattern: at the 6-month horizon, individuals with 4 *−* 6 years of history achieved the highest performance, whereas at the 36-month horizon, patients with *≥* 6 years of history consistently showed superior discrimination, suggesting that longer longitudinal records become increasingly important for earlier prediction of ADRD onset.

**Figure 4:**
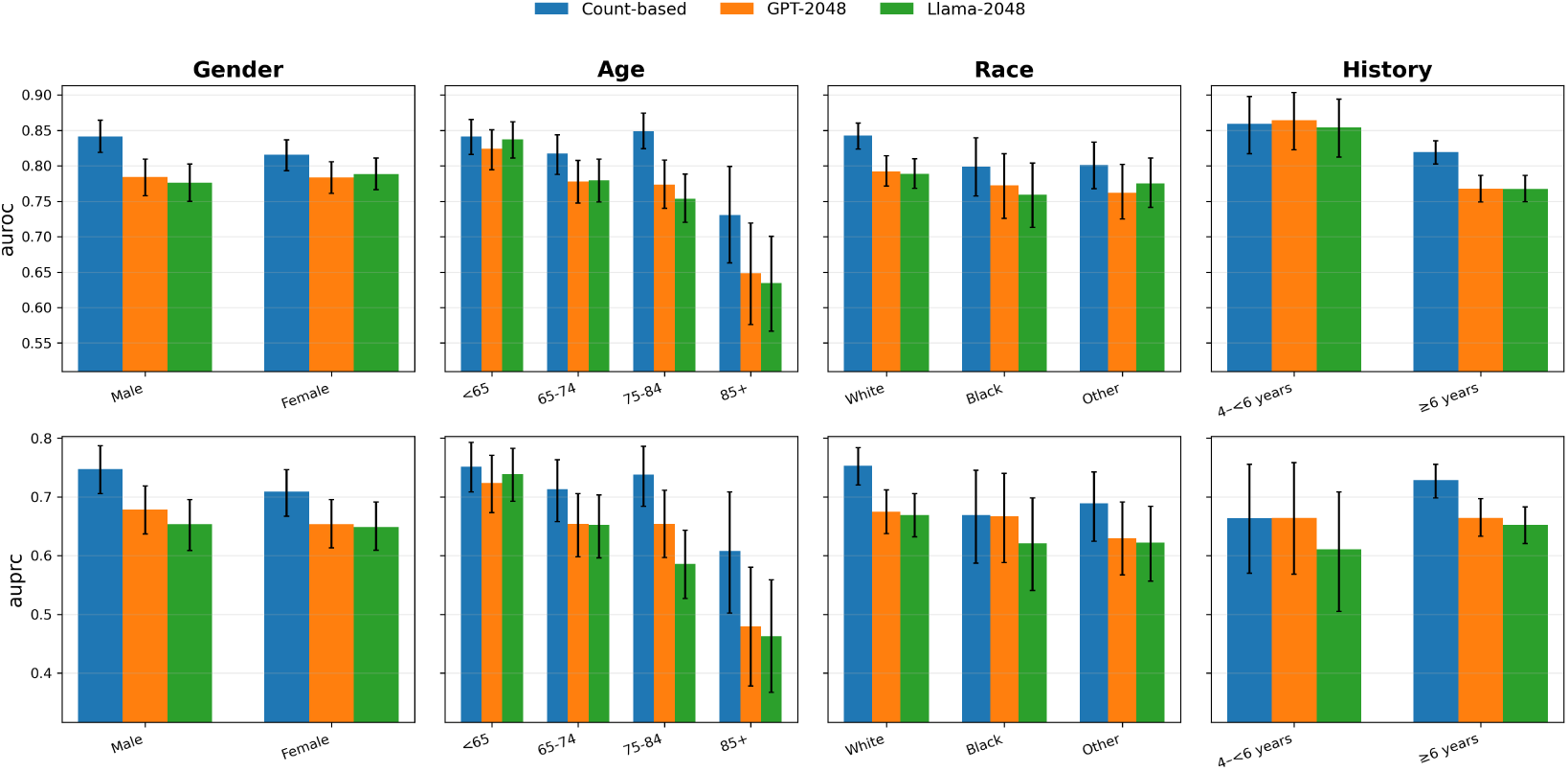
Performance across clinically relevant patient subgroups for AoU dataset. AUROC (top) and AUPRC (bottom) of the representative Count-based, GPT-2048, and LLaMA-2048 models on the held-out test set for the CD1 cohort at the 6-month prediction horizon, stratified by sex, age, race, and pre-prediction clinical history length. Error bars denote 95% bootstrap percentile intervals estimated within each subgroup.

## 3 Discussion

Early identification of individuals at risk for Alzheimer’s disease and related dementias remains a major challenge for population-scale clinical decision support. Using standardized longitudinal EHR data from two independent health systems, we systematically evaluated ADRD prediction across four clinically meaningful lead times and multiple cohort definitions while comparing interpretable count-based representations with publicly available pre-trained clinical foundation models. Across nearly all prediction horizons, count-based models achieved the best overall discrimination and calibration, whereas pretrained foundation-model embeddings remained competitive, particularly at longer prediction horizons where the performance gap narrowed. However, increasing the pretrained model context length from 512 to 2048 tokens produced only modest and architecture-dependent improvements, suggesting that larger context windows alone are insufficient to substantially improve downstream ADRD prediction using frozen pretrained representations.

Temporal feature-importance analysis demonstrated that the clinical information supporting prediction evolved with disease progression. At longer prediction horizons, models relied primarily on nonspecific indicators of healthcare utilization, including visit frequency, diagnosis frequency, observation frequency, and medication utilization, whereas disease-specific cognitive phenotypes, particularly amnesia and mild cognitive impairment, became increasingly important closer to diagnosis. Psychiatric disorders, laboratory measurements, smoking history, and socioeconomic characteristics contributed more modestly and varied across cohort definitions. Because SHAP quantifies model reliance rather than causal effects, these findings should be interpreted as predictive patterns learned from longitudinal EHRs rather than evidence of causal risk factors.

Incorporating survey-derived lifestyle and family health information provided only modest improvements over EHR-only models, largely because many participants lacked survey data before the prediction cutoff. Consequently, routinely collected longitudinal EHR data accounted for most of the predictive signal for ADRD onset, while survey variables provided complementary information when available. External zero-shot validation on the UChicago dataset demonstrated reduced performance across all model families, which is expected given the demographic differences and substantially shorter longitudinal clinical histories relative to the AoU dataset. Although pretrained foundation-model representations remained competitive in some settings, they did not consistently overcome cross-institutional distribution shift, highlighting the continued importance of institution-specific adaptation for reliable deployment.

Future work should investigate whether integrating longitudinal EHRs with complementary data modalities, including genetic risk factors, clinical notes, biomarkers, neuroimaging, and wearable data, can improve prediction during earlier preclinical stages. Richer temporal modeling of disease trajectories, rather than static patient-level representations, may further capture evolving patterns preceding ADRD onset. Finally, prospective multi-institutional validation and deployment studies will be essential to determine the clinical utility, robustness, and equitable implementation of ADRD risk prediction models across diverse healthcare systems.

## 4 Methods

### 4.1 Ethical Approval

The University of Chicago Institutional Review Board (IRB) approved the study (IRB Number: CIRB24-2050), and the Ethics Committee waived the need for written informed consent from participants because this study involved secondary analysis of de-identified data and posed minimal risk to participants.

### 4.2 Data Source, ADRD Phenotype and Case Definitions

We conducted a retrospective matched case-control study using two longitudinal electronic health record (EHR) databases: the National Institutes of Health All of Us (AoU) ( 633k patients as of 06/1/3036) Research Program Controlled Tier Version 8, and UChicago EHR ( 1.6M patients as of 07/01/2026). AoU is a national research program that integrates participant demographics, surveys, physical measurements, EHRs, biospecimens, and genomic data giving access to the registered and controlled tier researchers through a cloud-based workbench [8]. For the present study, only longitudinal EHR, demographic, and self-reported survey data from AoU were used. UChicago EHR provides only longitudinal EHR, demographic data. EHR data were harmonized to the Observational Medical Outcomes Partnership Common Data Model (OMOP) in both databases.

ADRD cases were identified using curated clinical concept sets representing Alzheimer disease, vascular dementia, Lewy body dementia, frontotemporal dementia, mixed dementia, and related dementia phenotypes, as well as exposure to medications commonly used for dementia treatment. Qualifying anti-dementia medications included donepezil, galantamine, memantine, and rivastigmine. Diagnosis concepts were identified using standard SNOMED concepts [9, 10, 11, 12, 13] and their mapped ICD-9-CM and ICD-10-CM source codes [14], while medication exposures were identified using corresponding RxNorm concepts [15]. Complete diagnosis and medication concept sets are provided in Supplementary Table S1.

Mild cognitive impairment (MCI) was not included in the ADRD outcome because it represents a clinically heterogeneous state with variable trajectories: some individuals progress to dementia, whereas others remain stable or revert to normal cognition. Restricting the outcome to ADRD diagnoses and ADRD-directed medication exposure provided a more specific target for predicting clinically recognized dementia onset.

We evaluated two rule-based ADRD case definitions to examine the effect of phenotype ascertainment on model performance. Case definition 1 (CD1; diagnosis or medication) was the broadest definition and included participants with either a qualifying ADRD diagnosis or exposure to a qualifying anti-dementia medication. For CD1, the ADRD index date was defined as the earliest qualifying diagnosis or medication exposure. The index criteria distribution for CD1 cohort is presented in Supplementary Table S2. Case definition 2 (CD2; secondary-confirmed) included participants whose first qualifying ADRD event was a diagnosis represented by an eligible ICD-or SNOMED-mapped concept followed by anti-dementia drug exposure. The index date was the first qualifying ADRD diagnosis (Supplementary Figure S1).

The two definitions were evaluated as alternative case-ascertainment strategies. CD2 represents a narrower phenotype derived from the broader CD1 cohort and was not intended to form an exhaustive partition of CD1. Consequently, some participants included in CD1, such as those identified initially through medication exposure or whose diagnosisâmedication sequence did not satisfy the CD2 criteria, were not included in CD2. The exact temporal logic for assigning participants to each case definition followed a prespecified cohort-generation algorithm. Although CD2 was expected to improve phenotypic specificity, it substantially reduced the number of eligible cases and therefore statistical power.

Using the broad ADRD phenotype (CD1), the initial case pool comprised 4,074 participants with at least one qualifying ADRD diagnosis or anti-dementia medication exposure. The initial control pool comprised 398,850 participants with no qualifying ADRD diagnosis and no qualifying anti-dementia medication exposure throughout their available EHR history. Initial cohort identification and case definitions are summarized in Figure 5.

**Figure 5:**
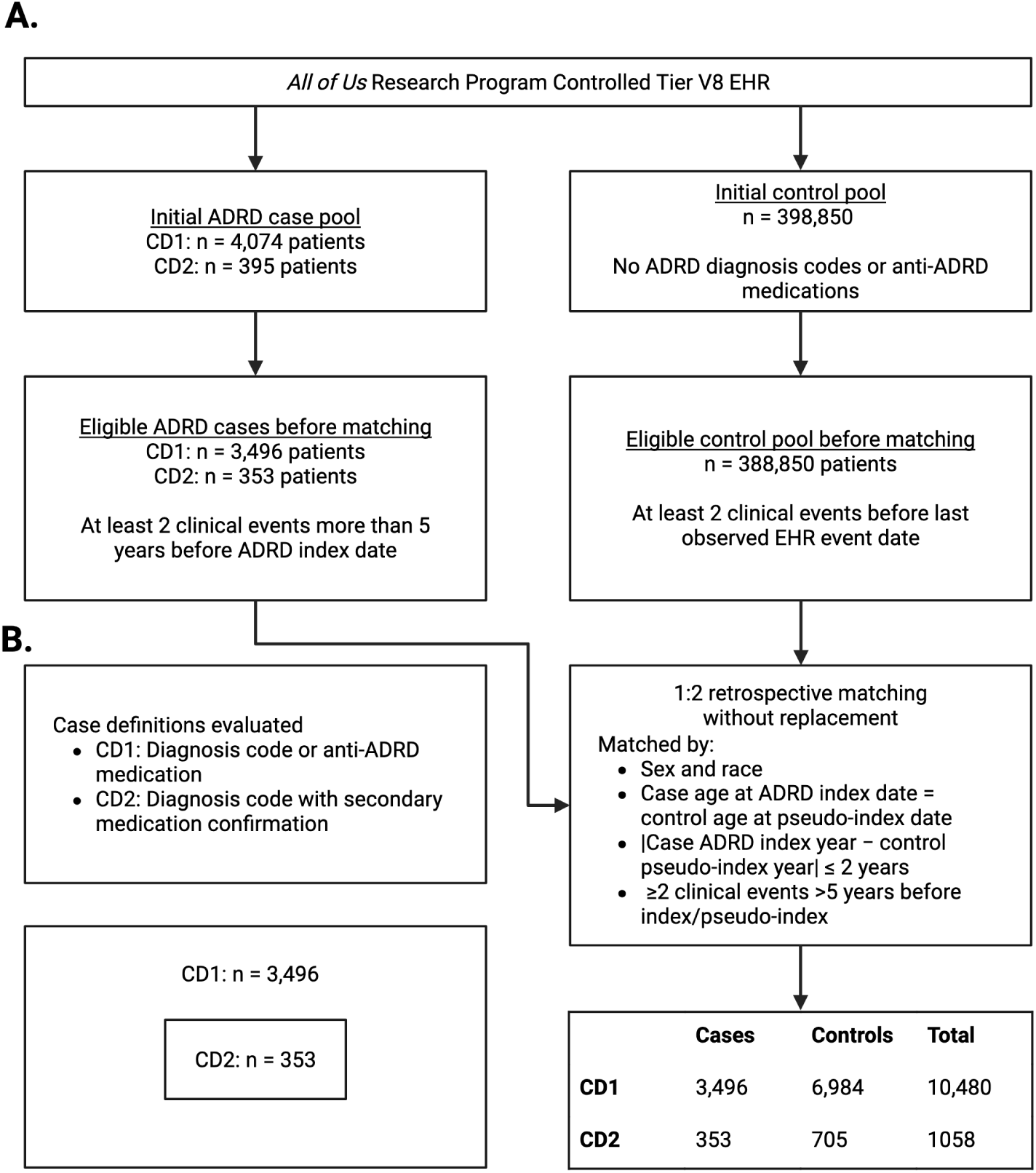
Construction of matched ADRD case–control cohorts across alternative phenotype definitions for AoU dataset. **(A)** Participants were screened from the AoU Controlled Tier V8 EHR population and evaluated for sufficient longitudinal history before cohort-specific retrospective matching. Matching was conducted separately for each ADRD definition, with control pseudo-index dates assigned to align age and calendar time while preserving the required pre-index observation period. **(B)** The two rule-based phenotypes represent distinct levels and sources of ADRD ascertainment: CD1 provides the broadest definition, while CD2 is the secondary-confirmed cohort. The resulting samples were used as separate analytic cohorts in subsequent prediction experiments.

### 4.3 Case-Control Cohort Construction and Matching

The initial case pool was required to have at least two clinical events more than five years before the case index date. Similarly, the initial control cohort was filtered to have at least two clinical events before the last recorded EHR event date. This criterion made the analytic cohorts more representative of individuals with sustained healthcare-system engagement than the broader AoU population. This resulted in the eligible pool of ADRD cases (n=3,496) and control (n=388,850) for final matching.

Each case was matched up to two controls without replacement. Candidate controls were restricted to participants with the same recorded sex and race as the corresponding case. For each candidate control, a pseudo-index date was calculated such that the controlâs age at pseudo-index equaled the matched caseâs age at ADRD index. To reduce temporal bias in retrospective case-control prediction studies by aligning the temporal context and longitudinal observation history between cases and controls, a control was considered eligible for matching only if: (1) the pseudo-index date fell within the control’s observed EHR interval; (2) the control had at least two clinical events recorded more than five years before the pseudo-index date, ensuring sufficient pre-index longitudinal history; and (3) the absolute difference between the case index date and the control pseudo-index date was no greater than two calendar years. These criteria were designed to align the prediction horizon and longitudinal observation history between matched cases and controls, consistent with recommendations for mitigating temporal bias in case-control prediction studies and with epidemiologic principles of risk-set sampling. [16, 17, 3]. Eligible cohort identification, matching process, and final analytic cohort sizes across case definitions for AoU database are summarized in Figure 5. For UChicago EHR database, we applied the same inclusion and exclusion criteria used for AoU database and identified 3,629 ADRD cases and 9,260 matched controls.

### 4.4 Observation Period and Prediction Horizons

For cases, the index date was defined according to the first qualifying event specified by the corresponding ADRD case definition. For controls, the pseudo-index date assigned during matching served as the corresponding reference date. We evaluated ADRD onset prediction at 6-, 12-, and 24 and 36-month horizons before the case index or control pseudo-index date. For each prediction horizon, the prediction cutoff was calculated by subtracting the specified horizon from the index or pseudo-index date.

The observation period extended from the participantâs first eligible recorded clinical event through the prediction cutoff. Only information recorded on or before the prediction cutoff was eligible for model input. Clinical events and other time-varying information recorded between the prediction cutoff and the index or pseudo-index date were withheld. This temporal censoring was intended to prevent models from learning directly from diagnostic evaluations, treatment initiation, or other healthcare activity occurring during the period immediately preceding clinically recognized ADRD onset.

The requirement for at least two clinical events more than five years before the index or pseudo-index date was used to establish sufficient longitudinal EHR history and was distinct from the prediction horizon. Only information available by the prediction cutoff was used to generate model inputs. Accordingly, any time-varying demographic, survey, or clinical variables were required to have been recorded on or before the relevant cutoff. Variables recorded after the cutoff were excluded from that prediction-horizon analysis. The temporal study design is illustrated in Figure 1 A.

### 4.5 Data Extraction, Preprocessing, and Feature Representation

#### 4.5.1 Clinical EHR Extraction and Preprocessing

Longitudinal clinical data were extracted from the OMOP condition_occurrence, drug_exposure, procedure_occurrence, measurement, and visit_occurrence domains. Domain-specific records were transformed into a common event representation containing the participant identifier, event date or datetime, standardized clinical concept, clinical domain, and, when available, recorded measurement values, units, and reference-range information. Events were ordered chronologically within each participantâs record and censored at the prediction cutoff for each prediction-horizon analysis.

Exact same-day duplicate records were collapsed only when all available identifying attributes were identical, including the standardized clinical concept, clinical domain, recorded value, unit, and reference-range information. Repeated occurrences on different dates were retained. Distinct same-day records were also retained, including events from different domains and measurements with different values, units, or reference ranges, because these represented separate clinical information rather than redundant database entries.

Clinical preprocessing included checks for unparseable or implausible dates, events occurring after the prediction cutoff, duplicated participants, invalid standardized concepts, and missing or nonfinite numeric values. Patient timelines were examined to confirm chronological ordering and the presence of usable clinical information before the prediction cutoff. Participants without any usable model-compatible history after preprocessing were identified before model fitting.

#### 4.5.2 Survey-data Extraction and Processing

Unlike longitudinal EHR data, which capture repeated clinical events over time, survey data in the AoU dataset consist of cross-sectional self-reported questionnaires completed by a subset of participants. UChicago dataset does not include any survey data. To prevent information leakage, only survey responses recorded on or before each participant’s prediction cutoff date were eligible for inclusion, and survey-derived features were generated independently for each prediction horizon and trainâtest split.

Survey variables were selected from two domains previously associated with ADRD risk identified from the existing literature [18, 19, 20, 21, 22]: lifestyle behaviors and personal and family health history. Rather than directly using individual questionnaire items, clinically related responses were curated and aggregated into composite variables to improve robustness and reduce redundancy. Lifestyle features captured smoking and alcohol exposure, including binary indicators of lifetime and current smoking or alcohol use, as well as quantitative measures such as cigarettes smoked per day, years of smoking, and daily alcohol consumption. Smoking-related binary variables were constructed by integrating responses from multiple tobacco-use questionnaires (e.g., cigarettes, cigars, hookah, smokeless tobacco, and electronic cigarettes). Personal and family health history features included self-reported mental health and substance use disorders, sleep apnea, social phobia, and family history of dementia or other neurodegenerative disorders. Composite indicators were similarly constructed for clinically related concepts, such as active social phobia, by combining responses regarding physician diagnosis, treatment, and medication use.

Survey responses indicating skipped questions, “prefer not to answer,” “don’t know,” or logically inapplicable responses were treated as missing. Because survey completion varied substantially across participants, explicit missingness indicator variables were created for each survey-derived feature, allowing the models to distinguish between the absence of a reported characteristic and the absence of survey information. When multiple responses to the same questionnaire item were available, the most recent response obtained before the participant’s prediction cutoff was retained.

Continuous survey variables were standardized together with other continuous model inputs during classifier training. In contrast to the longitudinal EHR features, which were represented as cumulative counts of clinical events occurring before the prediction time, survey variables were modeled as static participant-level attributes representing the most recent available pre-prediction self-reported status.

#### 4.5.3 Count-based Clinical Representation

The count-based clinical representation summarizes each participant’s longitudinal EHR history using interpretable features derived from the OMOP Common Data Model (CDM). Following established EHR count-featurization approaches [23, 24], patient representations were constructed independently for each prediction horizon using only information available before the participant’s prediction cutoff.

Clinical information was extracted from five OMOP CDM domains mentioned in 4.5.1. The resulting representation consisted of three groups of features: 1) **Clinical concept indicators**, representing the presence of individual OMOP concepts before the prediction time; 2) **Healthcare utilization features**, including the total number of recorded clinical events, the number of unique clinical concepts, and domain-specific event counts (e.g., diagnosis, medication, procedure, laboratory, and observation counts); 3) **Demographic features**, including sex, race, ethnicity, and age categorized into 5-year intervals.

To preserve temporal information while maintaining a fixed-length representation, clinical events were aggregated within four time windows defined relative to the prediction time: 0â24 hours, 1â7 days, 8â30 days, and >30 days before prediction. Features from each temporal window were concatenated to form the final longitudinal patient representation.

Continuous laboratory measurements were converted into binary abnormality indicators based on whether the recorded value exceeded or fell below the reference range documented in the EHR, rather than using raw numeric values.

To prevent information leakage, the feature vocabulary (i.e., the set of retained OMOP concepts and feature ordering) was derived exclusively from the training data. The learned vocabulary and column ordering were subsequently applied unchanged to the validation and held-out test sets. Consequently, concepts not observed during training were ignored during feature construction in the evaluation datasets.

#### 4.5.4 CLMBR-compatible Timelines and Representation Extraction

For the clinical foundation-model analyses, each participant’s longitudinal EHR history prior to the prediction cutoff was transformed into a chronological sequence of clinical events compatible with the Foundation Medical Record (FEMR) framework. Eligible events from the OMOP CDM were mapped to the standardized clinical vocabulary used by each pretrained CLMBR model [25, 26], while preserving their temporal ordering. Events occurring after the prediction cutoff were excluded to prevent information leakage. The complete pre-cutoff timeline was provided as input to the CLMBR tokenizer and encoder without manual truncation to a predefined context length. Sequence padding, truncation, and tokenization were performed internally by the pretrained CLMBR framework according to the model-specific context-length configuration. Consequently, each model automatically retained the appropriate portion of the longitudinal history consistent with its architecture.

Unlike the count-based representation, which summarizes clinical history using manually engineered features, the clinical foundation model represents the patient’s complete longitudinal record as a sequence of timestamped clinical tokens. Each participant’s sequence was tokenized using the fixed vocabulary distributed with the corresponding pretrained CLMBR model, without modifying or extending the tokenizer vocabulary during this study. Clinical concepts not recognized by the pretrained tokenizer were discarded prior to representation extraction.

A patient-level representation was obtained by applying the frozen pretrained CLMBR encoder to the pre-cutoff clinical sequence. For each participant and prediction horizon, the encoder produced a fixed-length 768-dimensional embedding summarizing the entire longitudinal clinical history available up to the prediction cutoff. These embeddings were used directly as input features for downstream classifiers, while the CLMBR encoder remained frozen throughout model development. Each pretrained model retained its native maximum context length. Sequences shorter than the maximum context length were left-padded, whereas longer sequences were truncated to retain the most recent eligible clinical events.

Representation-level quality control verified that exactly one embedding was generated for every participant and prediction horizon, with the expected embedding dimensionality, no missing or non-finite values, and complete consistency among participant identifiers, case-control labels, matched-set identifiers, and train-validation-test assignments.

### 4.6 Data Splitting and Leakage Prevention

Model training and evaluation were conducted separately for each ADRD case definition. Each matched case-control cohort was divided into a 70% training set and a 30% held-out test set. Splitting was performed at the matched-set level rather than at the individual-participant level. Each case and all of its matched controls shared a common matched-set identifier and were assigned together to either the training or test partition. This prevented a case from appearing in one partition while demographically and temporally matched controls appeared in another.

Within each case definition, the trainâtest split was designed to preserve the distribution of the outcome and key demographic characteristics. Stratified sampling was performed using sex, race, 5-year index-age categories, and the ADRD ascertainment criterion (first qualifying diagnosis concept or first qualifying anti-dementia medication concept), with sparse strata collapsed when necessary. The resulting train and test assignments were fixed and reused across all feature representations and downstream classifiers, ensuring fair model comparisons on identical participants.

Within the 70% training set, group-aware three-fold cross-validation was used for hyperparameter tuning, and model selection. Matched sets remained intact within each cross-validation fold. No matched set was permitted to contribute participants to both the training and validation portions of a fold.

All learned preprocessing and feature-construction procedures were restricted to the training data. These included clinical vocabulary construction, rare-concept filtering, regularization tuning, and class-weight selection. During cross-validation, procedures that could influence model selection were fitted using the corresponding inner-training folds and applied to the associated validation fold. The held-out test set was not used to construct features, select hyperparameters, choose decision thresholds, determine training duration, or select model configurations.

After the final configuration was selected, its preprocessing procedures and model parameters were refitted using the complete training set. The resulting model was then evaluated once on the held-out test set. Split quality-control analyses verified that no participant identifier or matched-set identifier overlapped between the training and test sets or between the training and validation portions of each cross-validation fold. Each partition was also checked to confirm the presence of both cases and controls and preservation of the approximate case-control matching ratio.

### 4.7 Model Development and Evaluation

We evaluated two primary approaches to represent longitudinal EHR information: sparse count-based clinical representations and dense representations generated using pretrained CLMBR clinical foundation models. We evaluated pretrained CLMBR models with four backbone architectures, including Transformer-base, LLaMA-style, GPT-style, and Mamba sequence models. For the LLaMA-style and GPT-style CLMBR architectures, we extracted embeddings from pretrained models with varied context length (512 and 2048) to assess whether retaining a larger number of preceding clinical events improved prediction performance. Sparse clinical-concept count vectors and CLMBR derived embeddings were used to train logistic regression (with L2 [27] and elasticnet [28] regularization) [29], XGBoost [30], and LightGBM [31] models. Logistic-regression hyperparameters included regularization strength, class weight, L1 ratio (for elasticnet). Hyperparameters for XGBoost and Light-GBM included max_depth (XGBoost), num_leaves (LightGBM), learning_rate. Hyperparameters were selected using group-aware three-fold cross-validation within the development set.

Survey-derived variables were concatenated with the longitudinal EHR count-based features and the foundation model embeddings using early fusion strategy to form a unified feature representations for downstream classification. Rather than learning latent survey representations using a dedicated neural encoder, survey variables were incorporated directly as engineered features. Given the relatively small number of survey variables, substantial missingness, and the study’s emphasis on interpretable multimodal integration, early feature fusion provides a simpler and more robust modeling strategy while reducing the risk of overfitting. The count-based model and CLMBR models where modeled with and without different variants of survey features (e.g. smoking, alcohol, survey (all survey features combined that are mentioned in 4.5.2)). These comparisons were used to assess whether participant-reported information improved prediction beyond longitudinal structured EHR information alone.

#### 4.7.1 Experimental Setup and Model Selection

Count-based logistic regression and XGBoost were used as classical baselines for each case definition and prediction horizon. CLMBR experiments assessed whether pretrained sequential EHR representations improved performance relative to sparse clinical-concept counts. Additional analyses compared backbone architectures and context lengths. Model selection was performed exclusively within the training set using 3-fold cross-validation. For each variant of the model (with or without survey data and the varying context length of the CLMBR models), the configuration was selected that maximizes the AUROC stability score ^1^ across folds. The selected model was then retrained on the full training set and evaluated once on the independent held-out test set.

#### 4.7.2 Performance Evaluation

Discrimination was evaluated using the area under the receiver operating characteristic curve and area under the precision-recall curve. Probabilistic prediction accuracy was evaluated using the Brier score. Threshold-based performance measures included sensitivity, specificity, precision, F1 score, and balanced accuracy. AUROC, AUPRC, and Brier score were calculated from continuous predicted probabilities and did not depend on a classification threshold.

During development, empirically selected decision thresholds generally ranged from 0.30 to 0.50. To standardize comparisons across model variants, case definitions, and prediction horizons, a fixed probability threshold of 0.50 was used for primary threshold-based evaluation on the held-out test set.

### 4.8 Top Feature Interpretation

To improve the interpretability of the count-based models, we quantified feature importance using SHAP (SHapley Additive exPlanations) [32] values computed from the final XGBoost models. Although LightGBM achieved the highest predictive performance among the count-based classifiers, followed closely by XGBoost, XGBoost was selected for interpretation because it provides robust and widely adopted TreeSHAP explanations with well-established support for feature-level attribution in tree-based models. For each cohort definition and prediction horizon, TreeSHAP values were computed from the final retrained XGBoost model on the held-out test set. Feature importance was quantified as the mean absolute SHAP value across all patients, reflecting the average contribution of each feature to model predictions irrespective of direction. Features were ranked independently within each cohort definition and prediction horizon according to their mean absolute SHAP values, and the top 20 features from each model were retained for downstream comparison. To facilitate visualization across cohort definitions and prediction horizons, the union of all selected top-ranked features was assembled into a combined heatmap. Heatmap values represent the mean absolute SHAP value for each feature within each model. Features that did not rank among the top 20 for a given cohort definition and prediction horizon were assigned a value of 0.0 in the heatmap, indicating that they were not selected among the highest-contributing features in that model rather than having zero predictive contribution. To better interpret the health-care utilization features identified by the global SHAP analysis, we performed secondary analyses on the held-out test of a selected cohort for the highest-performing count-based XGBoost models. For the visit frequency feature, patients were stratified into high-and low-SHAP groups using the median SHAP value of the feature. Within each group, we quantified the prevalence of healthcare encounters across OMOP visit domains, including inpatient, outpatient, emergency department, home health, and other visit types. Visit-type prevalence was calculated as the proportion of patients with at least one encounter of each type before the prediction cutoff, allowing us to determine which encounter settings primarily contributed to elevated visit frequency SHAP attribution. To further characterize the global drug utilization feature, we mapped individual RxNorm ingredient concepts to higher-level therapeutic drug classes using the Anatomical Therapeutic Chemical (ATC) classification hierarchy. Major therapeutic classes were selected based on overall frequency within the study population. Patients were similarly stratified into high-and low-SHAP groups according to the median SHAP value for the drug utilization feature. For each patient, the contribution of each therapeutic class was quantified as the proportion of all pre-cutoff medication exposures belonging to that class, thereby normalizing for differences in overall medication burden. Therapeutic-class distributions were then compared between the two SHAP groups to identify medication categories associated with positive or negative model attribution while accounting for variation in total drug utilization.

#### 4.8.1 Subgroup Evaluation

For the selected model, performance was evaluated on the held-out test set within prespecified subgroups defined by sex, race, age at prediction, and clinical history length. The same trained model was applied to each subgroup without subgroup-specific retraining. Age was calculated at the prediction cutoff and categorized as younger than 65 years, 65â74 years, 75â84 years, and 85 years or older. Pre-prediction clinical history length was categorized as 4â6 years and *≥* 6 years of observable EHR history. To illustrate the variability of subgroup-specific performance estimates, we computed bootstrap percentile intervals by resampling participants within each subgroup with replacement. The original matched case-control structure was not preserved during resampling; therefore, these intervals should be interpreted as descriptive measures of sampling variability rather than formal inferential confidence intervals for comparing subgroup performance.

### 4.9 Computing Platform

Data analysis was conducted on a Unix system equipped with 8 CPU having 4 cores (IntelÂ® XeonÂ® CPU @ 2.30GHz) and 50 GB of memory. Data curation, visualization, and modeling were carried out using Python 3.10.19 with packages scikit-learn 1.6.1, seaborn 0.13.2, matplotlib 3.10.1, PyTorch 2.0.1+cu118, transformers 5.16.1.

## 5 Limitations

This study has several limitations. First, our primary case definition (CD1) used a broad ADRD phenotype that included individuals identified through ADRD diagnosis code or anti-dementia medication exposure. Because these medications may occasionally be prescribed for conditions other than ADRD, some outcome misclassification is possible. To address this concern, we additionally evaluated a more specific secondary-confirmed case definition (CD2), in which ADRD diagnosis preceded anti-dementia medication use, and observed consistent overall trends across both case definitions. Second, our SHAP analyses were designed to characterize global temporal changes in model reliance across prediction horizons and cohort definitions rather than sex-specific predictor differences. Consequently, we did not perform sex-stratified feature attribution analyses, although such analyses may provide additional insights into potential biological or clinical heterogeneity. Third, SHAP values quantify feature contributions to model predictions rather than biological mechanisms or causal pathways; therefore, the reported feature importance should be interpreted as model reliance within longitudinal EHR data rather than evidence of disease etiology. Finally, interpretability analyses were limited to the count-based models. We did not investigate the internal representations learned by the pretrained clinical foundation models, and understanding the clinical information encoded within these embedding spaces remains an important direction for future research.

## 6 Data Availability

The data used in this study contain sensitive patient information and therefore are not publicly available. Access to data is restricted to protect patient privacy and confidentiality. Researchers interested in accessing the data for academic purposes can contact the corresponding author for more information on the terms and conditions of access to the data.

## 7 Code Availability

The code for the modeling approach can be found in https://github.com/treena908/ADRD_onset_prediction

## 8 Acknowledgements

This research is funded by grant R56AG082167 from the National Institute on Aging, of the National Institutes of Health (NIH), OT2 OD036445 from All of Us Research Program, NIH, P30 ES027792 from National Institute of Environmental Health Sciences.

## 9 Author contributions

S.F.: Study design, NLP-related data curation, model design, model interpretability assessment, data analysis, manuscript writing. A.A.: Study design, NLP-related data curation, model interpretability assessment, manuscript writing, T.R., M.D.: Expert opinion from a clinician’s perspective, model interpretability assessment, manuscript revision, H.A.: Study design, Expert opinion on cohort selection, manuscript revision.

## Supplementary Material

**Figure S1:**
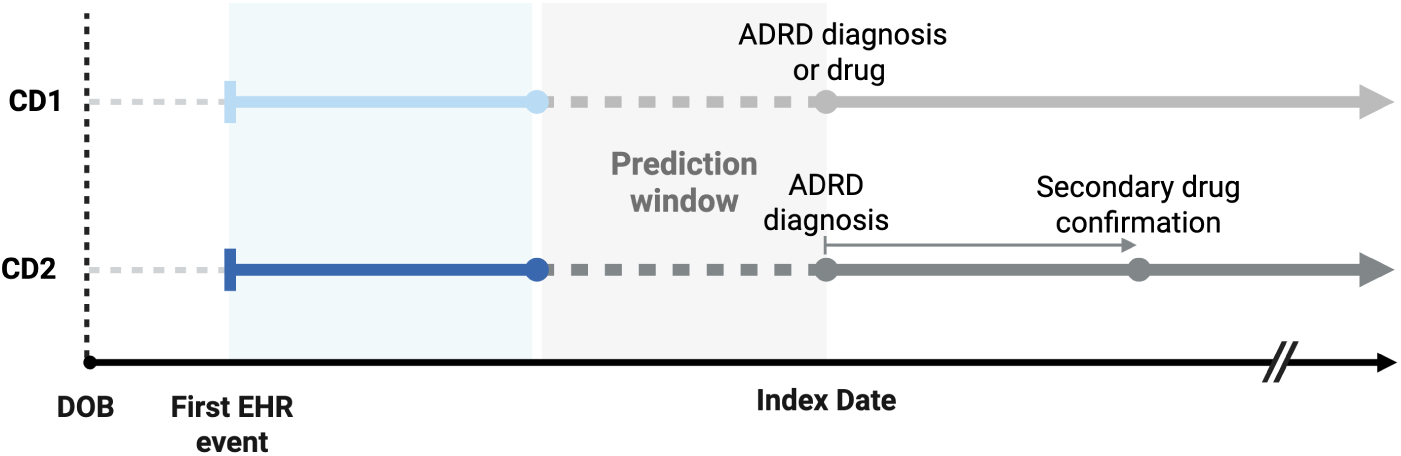
Schematic illustration of the longitudinal relationship between EHR observation, prediction, and ADRD case ascertainment for the primary case definitions. For CD1, the index date was defined by the earliest qualifying ADRD diagnosis or anti-ADRD medication exposure. For CD2, the index date corresponded to the qualifying ADRD diagnosis, with a subsequent anti-ADRD medication exposure serving as secondary confirmation. For each prediction horizon, the prediction window immediately preceded the index date, and only EHR information recorded before the start of that window was available for model input. DOB, date of birth; EHR, electronic health record; ADRD, Alzheimerâs disease and related dementias.

**Figure S2:**
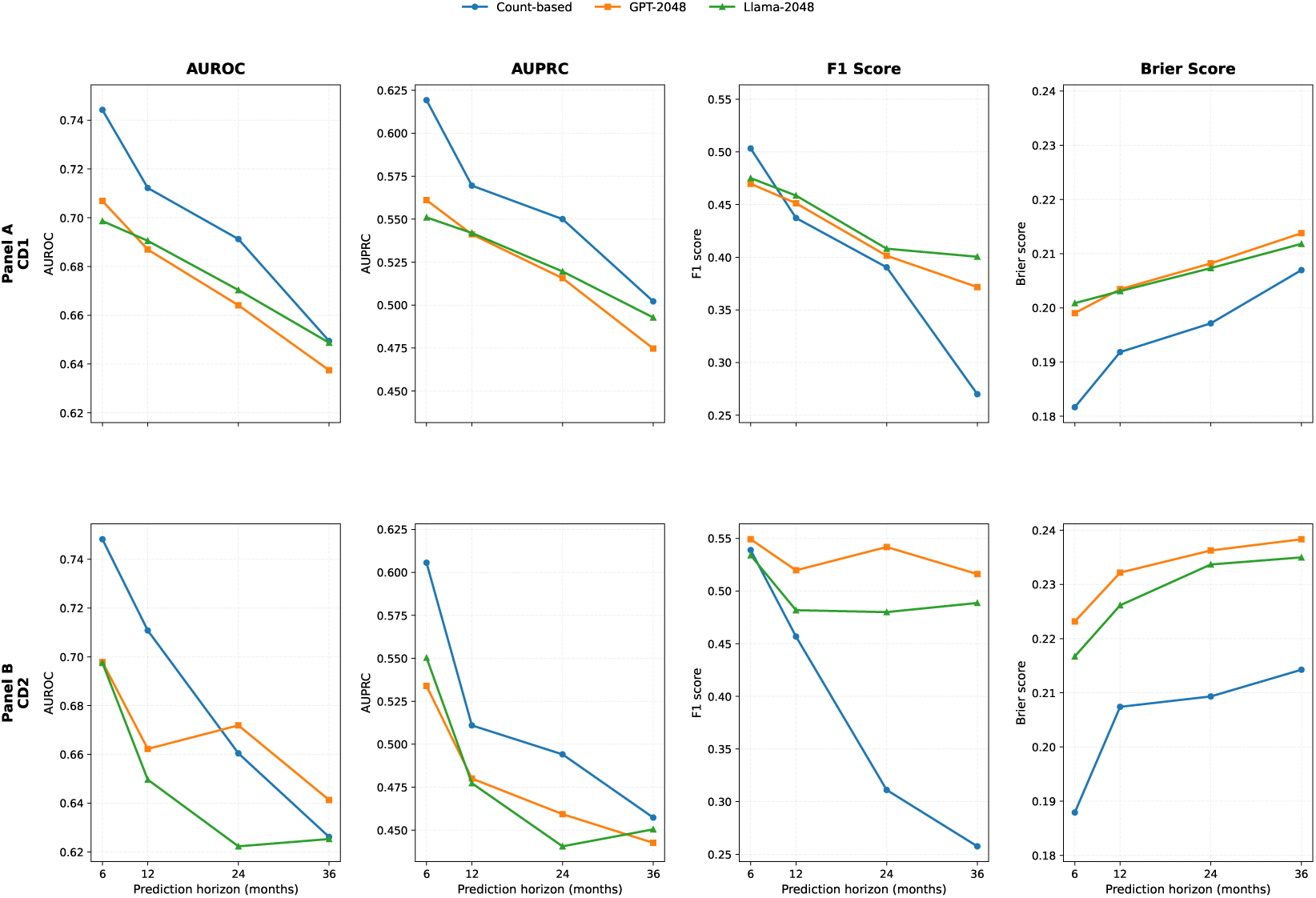
Held-out test set results on UChicago EHRs trained on UChicago train set.

**Figure S3:**
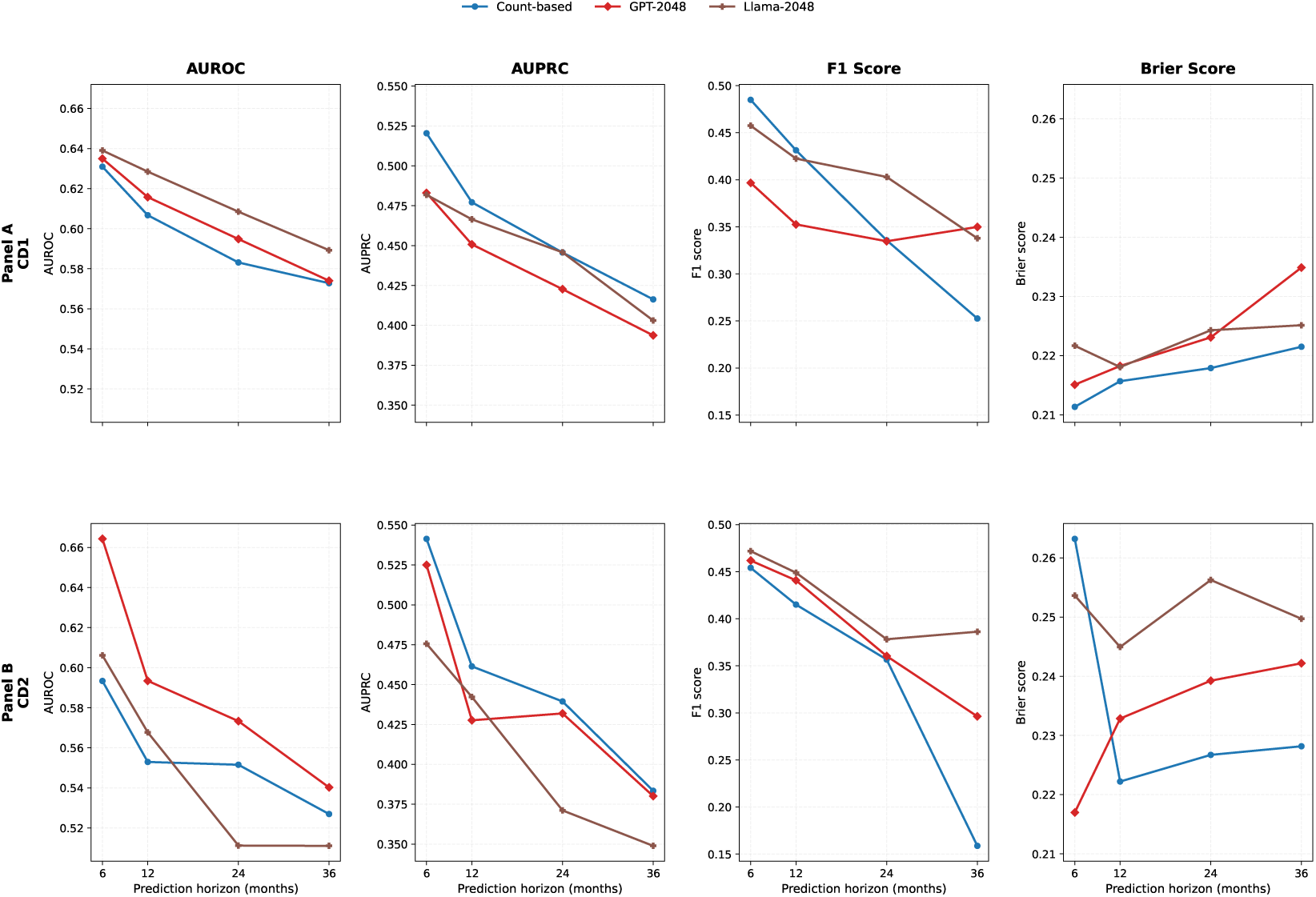
Held-out test set results on UChicago EHRs with zero-shot transfer from AoU train set.

**Figure S4:**
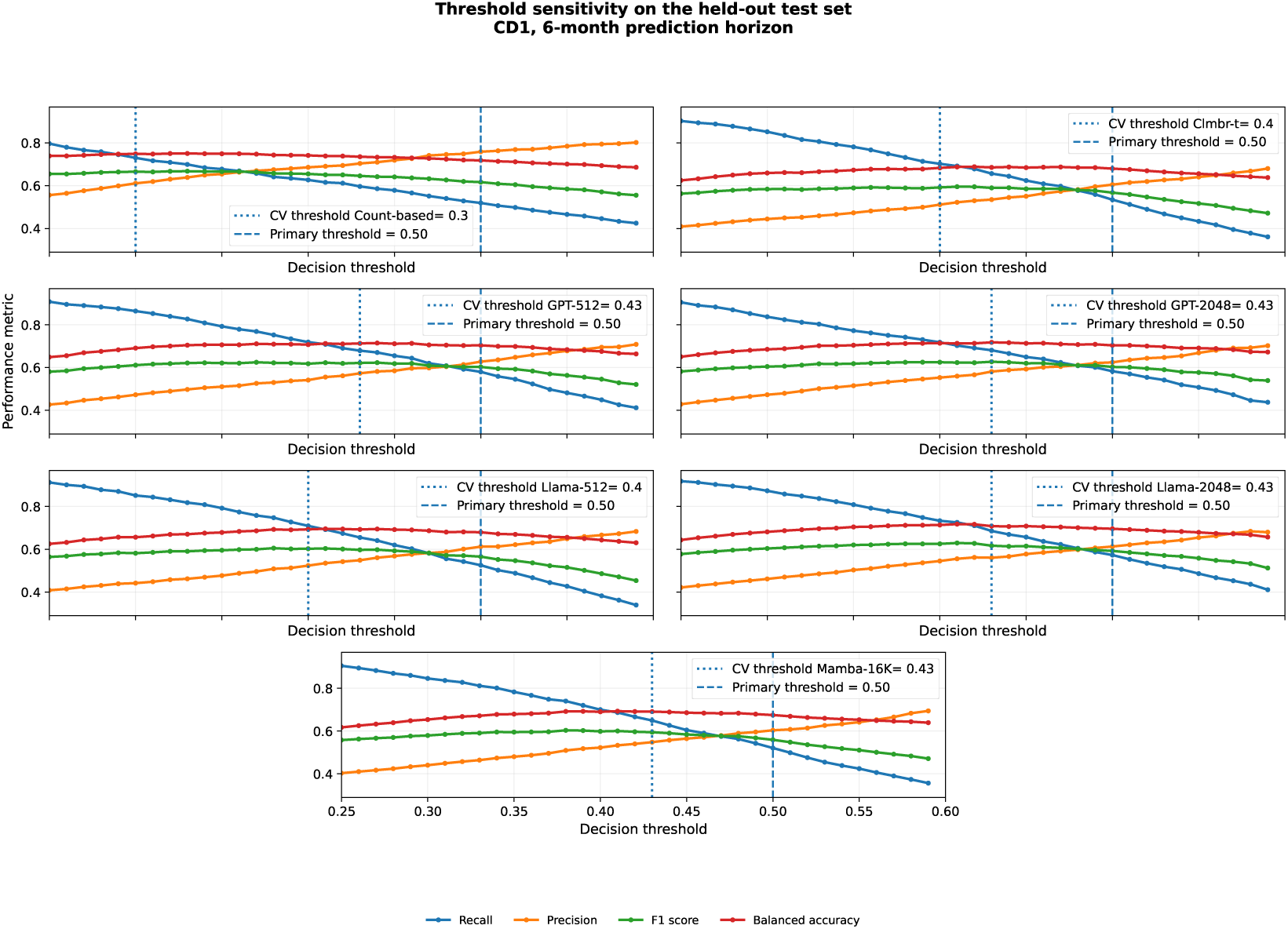
Threshold sensitivity analysis as identified from the held-out test-set results of AoU dataset, in the CD1 cohort at the 6-month prediction horizon. Recall, precision, F1 score, and balanced accuracy were evaluated across decision thresholds from 0.25 to 0.60 for the best-performing representative models from the count-based and CLMBR model families. Dashed vertical lines indicate the fixed threshold of 0.50 used in the primary test-set analysis, whereas dotted vertical lines indicate the model-specific thresholds selected during cross-validation. Increasing the decision threshold improved precision but reduced recall, while F1 score and balanced accuracy remained comparatively stable across intermediate thresholds.

**Table S1:** OMOP standard concepts used to define ADRD cases.

| Type | Concept IDs/ICD-10 code (Name) |
| --- | --- |
| Conditions | 443605/ICD-10/F01 (Vascular dementia), 380701/ICD-10/G31.83 (Diffuse Lewy body disease), 4196433/ICD-10/G31.83 (Senile dementia of the Lewy body type), 43021816/ICD-10/G30.8, ICD-10/F00.2 (Mixed dementia), 37109056/ICD-10/F01.50 (Vascular dementia without behavioral disturbance), 4043378/ICD-10/G31.09 (Frontotemporal dementia), 44782763/ICD-10/G31.83 (Lewy body dementia with behavioral disturbance), 378419/ICD-10/G30.9 (Alzheimer’s disease), 37018688/ICD-10/F01.51 (Vascular dementia with behavioral disturbance). |
| Medications | 715997 (Donepezil), 733523 (Rivastigmine), 757627 (Galantamine), 701322 (Memantine). |

**Table S2:** Index criteria distribution.

| Index Criteria | Distribution (%) |  |
| --- | --- | --- |
|  | AoU | UChicago |
| Anti-ADRD medication | 63.0% | 52.2% |
| Alzheimer’s disease | 18.0% | 20.3% |
| Vascular dementia | 14.4% | 24.9% |
| Frontotemporal dementia | 2.5% | 1.5% |
| Lewy body dementia | 2.1% | 1.2% |
| Mixed dementia | 0.1% | - |

**Figure S5:**
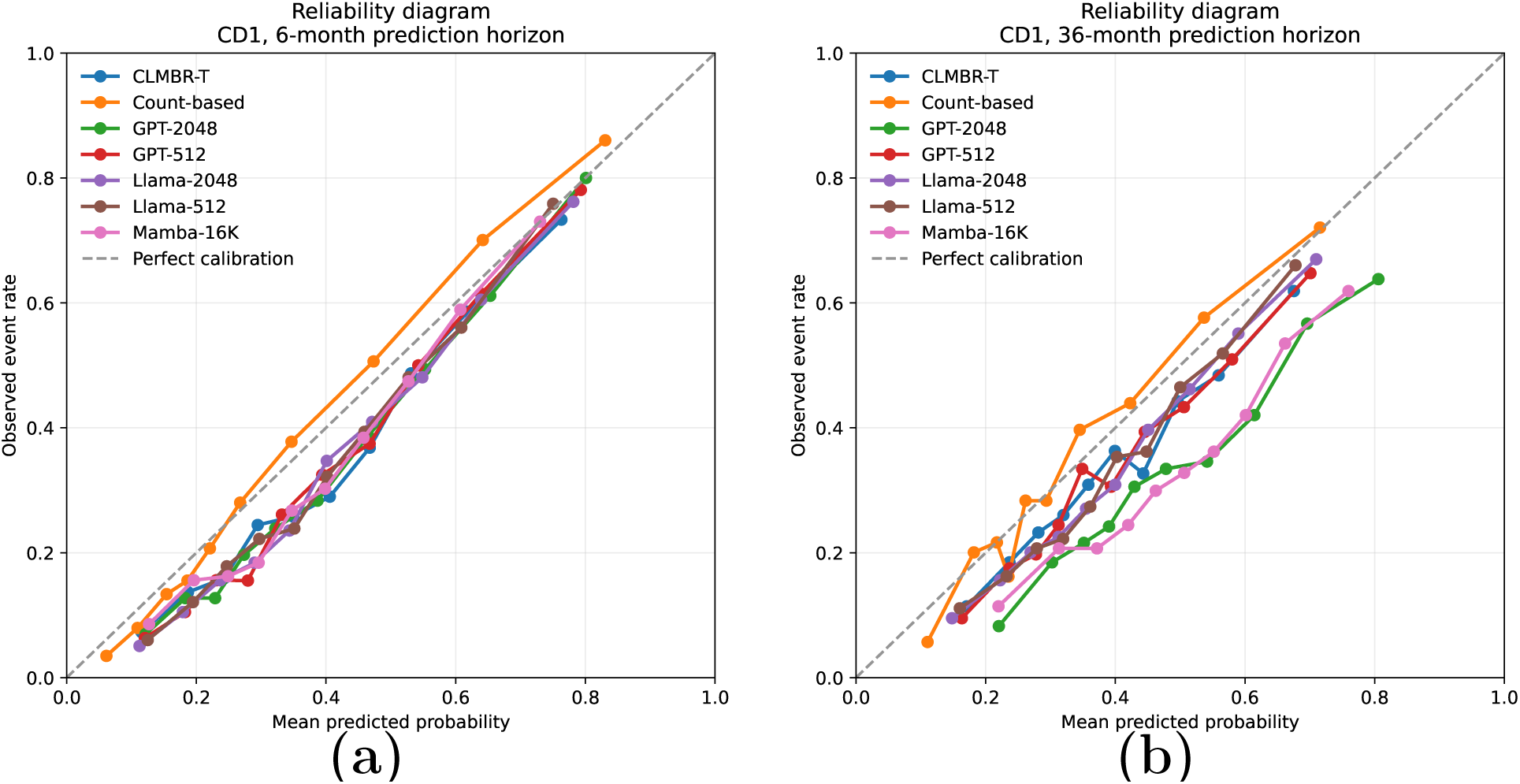
Reliability curve as identified from held-out test set of AoU dataset for CD1 cohort at (a) 6-month prediction horizon (b) 36-month prediction horizon.

**Table S3:**
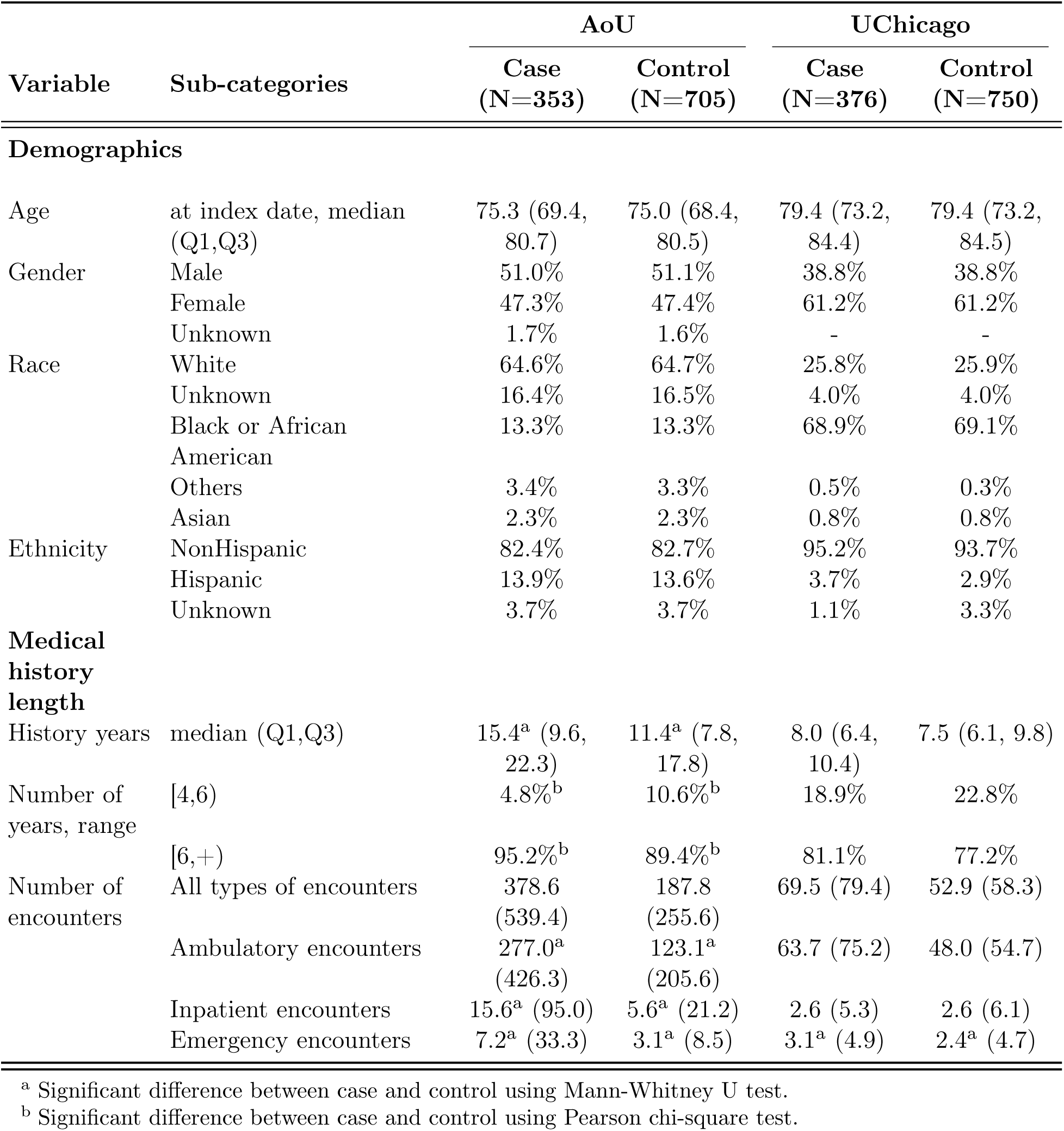
Demographic characteristics, healthcare utilization, and EHR history of ADRD cases and matched controls for CD2 cohort.

**Table S4:** Cross-validation performance of the best-performing feature variant within each model family for CD1 across all prediction horizons.

| Prediction horizon | Model family | Selected variant | Classifier | AUROC | AUPRC | F1 score | Brier score |
| --- | --- | --- | --- | --- | --- | --- | --- |
| 6 | Count-based | Count+Smoking | LightGBM | $0.806 \pm 0.000$ | $0.690 \pm 0.009$ | $0.644 \pm 0.011$ | $0.162 \pm 0.002$ |
| | CLMBR-T | Emb+Smoking | LR | $0.748 \pm 0.016$ | $0.609 \pm 0.023$ | $0.592 \pm 0.017$ | $0.190 \pm 0.004$ |
| | GPT-512 | Emb+Smoking | LR | $0.769 \pm 0.009$ | $0.637 \pm 0.017$ | $0.613 \pm 0.011$ | $0.183 \pm 0.003$ |
| | GPT-2048 | Emb+Smoking | LR | $0.765 \pm 0.015$ | $0.631 \pm 0.028$ | $0.603 \pm 0.018$ | $0.184 \pm 0.005$ |
| | Llama-512 | Emb+Smoking | LR | $0.736 \pm 0.012$ | $0.585 \pm 0.016$ | $0.583 \pm 0.012$ | $0.195 \pm 0.003$ |
| | Llama-2048 | Emb+Smoking | LR | $0.759 \pm 0.013$ | $0.620 \pm 0.018$ | $0.602 \pm 0.010$ | $0.187 \pm 0.004$ |
| | Mamba-16K | Emb+Smoking | LR | $0.742 \pm 0.014$ | $0.603 \pm 0.027$ | $0.580 \pm 0.004$ | $0.192 \pm 0.004$ |
| 12 | Count-based | Count | LightGBM | $0.772 \pm 0.003$ | $0.640 \pm 0.010$ | $0.608 \pm 0.012$ | $0.175 \pm 0.002$ |
| | CLMBR-T | Emb+Lifestyle | LR | $0.723 \pm 0.012$ | $0.568 \pm 0.021$ | $0.561 \pm 0.029$ | $0.198 \pm 0.004$ |
| | GPT-512 | Emb+Lifestyle | LR | $0.737 \pm 0.012$ | $0.587 \pm 0.022$ | $0.581 \pm 0.009$ | $0.194 \pm 0.004$ |
| | GPT-2048 | Emb+Lifestyle | LR | $0.738 \pm 0.013$ | $0.590 \pm 0.016$ | $0.578 \pm 0.028$ | $0.193 \pm 0.004$ |
| | Llama-512 | Emb+Smoking | LR | $0.717 \pm 0.015$ | $0.555 \pm 0.016$ | $0.567 \pm 0.010$ | $0.200 \pm 0.005$ |
| | Llama-2048 | Emb+Smoking | LR | $0.740 \pm 0.013$ | $0.588 \pm 0.015$ | $0.591 \pm 0.015$ | $0.193 \pm 0.004$ |
| | Mamba-16K | Emb+Smoking | LR | $0.723 \pm 0.007$ | $0.577 \pm 0.017$ | $0.571 \pm 0.004$ | $0.197 \pm 0.002$ |
| 24 | Count-based | Count+Alcohol | LightGBM | $0.738 \pm 0.003$ | $0.597 \pm 0.005$ | $0.582 \pm 0.002$ | $0.186 \pm 0.000$ |
| | CLMBR-T | Emb+Lifestyle | LR | $0.682 \pm 0.015$ | $0.520 \pm 0.019$ | $0.530 \pm 0.014$ | $0.226 \pm 0.004$ |
| | GPT-512 | Emb+Lifestyle | LR | $0.702 \pm 0.017$ | $0.539 \pm 0.023$ | $0.553 \pm 0.018$ | $0.221 \pm 0.004$ |
| | GPT-2048 | Emb+Smoking | LR | $0.711 \pm 0.015$ | $0.558 \pm 0.021$ | $0.559 \pm 0.016$ | $0.200 \pm 0.003$ |
| | Llama-512 | Emb+Lifestyle | LR | $0.691 \pm 0.010$ | $0.522 \pm 0.009$ | $0.549 \pm 0.008$ | $0.206 \pm 0.003$ |
| | Llama-2048 | Emb+Smoking | LR | $0.713 \pm 0.014$ | $0.552 \pm 0.017$ | $0.559 \pm 0.016$ | $0.201 \pm 0.004$ |
| | Mamba-16K | Emb+Lifestyle | LR | $0.692 \pm 0.015$ | $0.528 \pm 0.020$ | $0.549 \pm 0.009$ | $0.206 \pm 0.003$ |
| 36 | Count-based | Count+Smoking | LightGBM | $0.710 \pm 0.004$ | $0.557 \pm 0.010$ | $0.546 \pm 0.022$ | $0.194 \pm 0.002$ |
| | CLMBR-T | Emb+Lifestyle | LR | $0.663 \pm 0.010$ | $0.498 \pm 0.016$ | $0.504 \pm 0.043$ | $0.212 \pm 0.002$ |
| | GPT-512 | Emb+Lifestyle | LR | $0.692 \pm 0.013$ | $0.521 \pm 0.021$ | $0.545 \pm 0.012$ | $0.206 \pm 0.003$ |
| | GPT-2048 | Emb+Smoking | LR | $0.696 \pm 0.009$ | $0.531 \pm 0.006$ | $0.536 \pm 0.007$ | $0.222 \pm 0.001$ |
| | Llama-512 | Emb+Smoking | LR | $0.677 \pm 0.006$ | $0.509 \pm 0.012$ | $0.522 \pm 0.045$ | $0.209 \pm 0.002$ |
| | Llama-2048 | Emb+Lifestyle | LR | $0.694 \pm 0.008$ | $0.530 \pm 0.011$ | $0.548 \pm 0.006$ | $0.205 \pm 0.002$ |
| | Mamba-16K | Emb+Lifestyle | LR | $0.669 \pm 0.014$ | $0.498 \pm 0.021$ | $0.529 \pm 0.017$ | $0.211 \pm 0.002$ |

Values are reported as mean ± standard deviation across three cross-validation folds. Within prediction horizon, and model family, only the feature variant selected according to the prespecified cross-validation model-selection procedure is shown. Lower Brier scores indicate better probabilistic accuracy.

**Figure S6:**
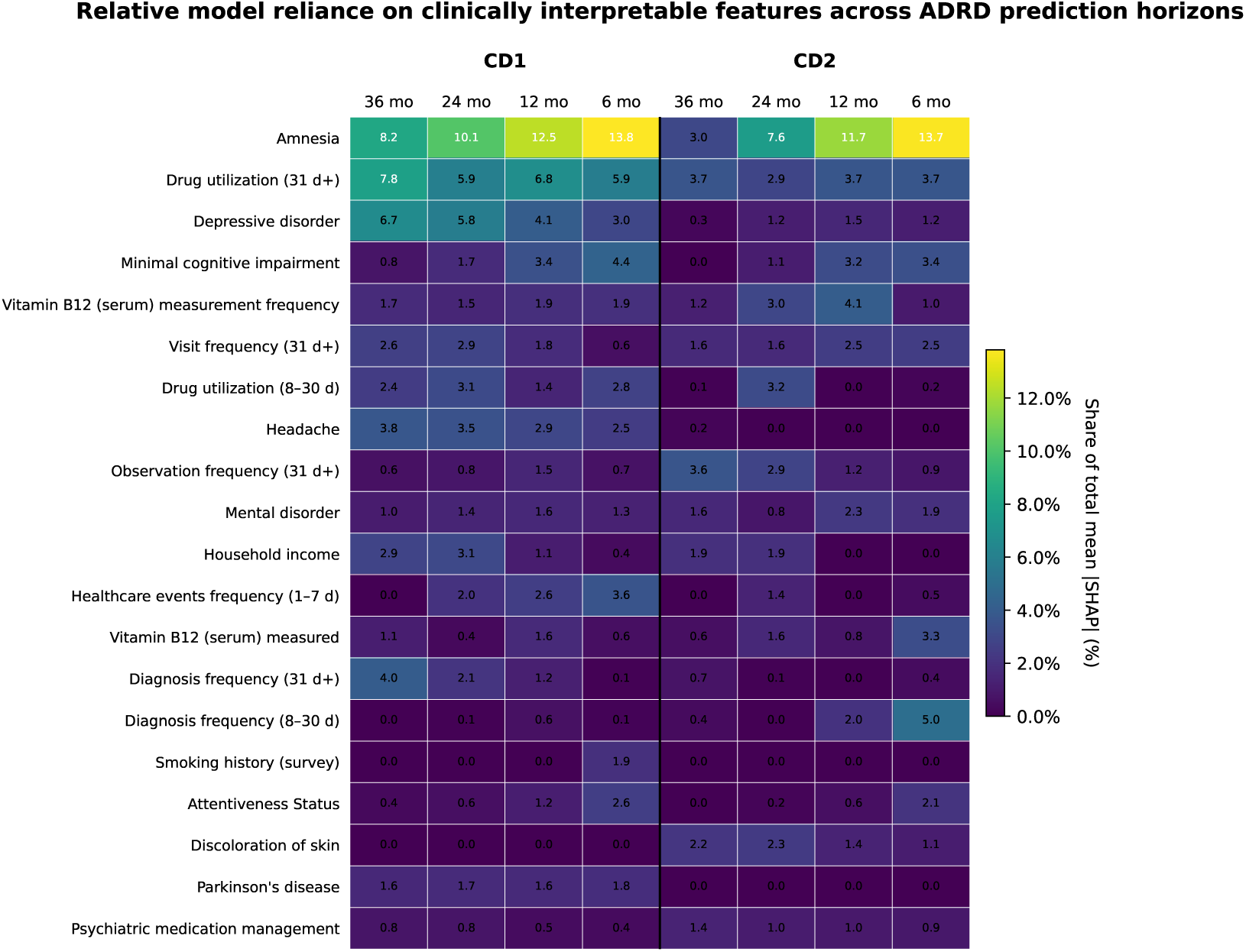
Cross-horizon feature importance for the top 20 predictors.

**Figure S7:**
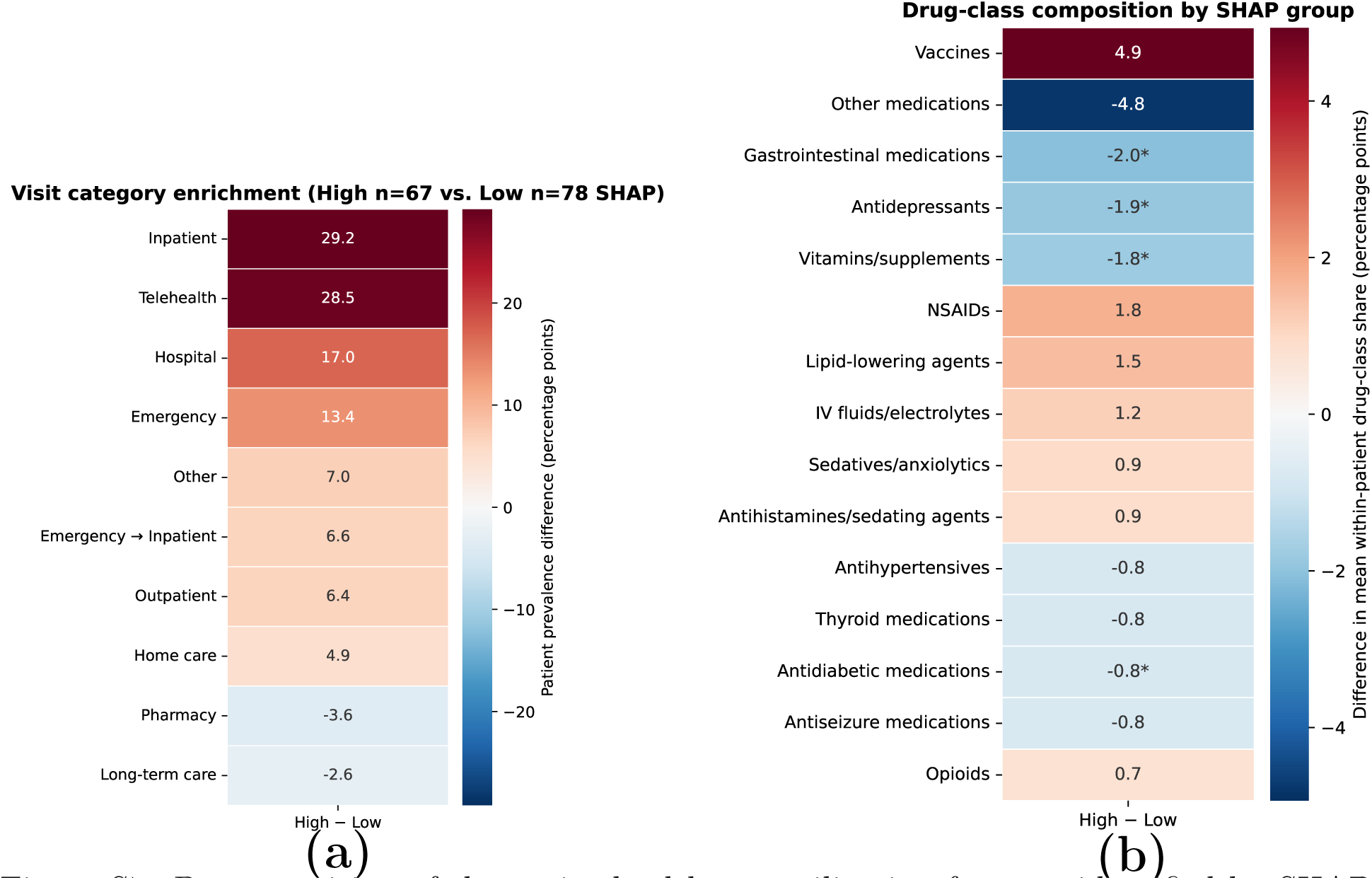
Decomposition of the major healthcare utilization features identified by SHAP. (a) Difference in the prevalence of encounter types between patients with high and low SHAP values for the visit-frequency feature. Values represent absolute percentage-point differences in the proportion of patients with at least one encounter of each visit type before the prediction cutoff. (b) Difference in normalized therapeutic-class composition between high- and low-SHAP groups (high *n* = 78 vs. low *n* = 43) for the drug-utilization feature. Values represent percentage-point differences in the mean within-patient proportion of medication exposures belonging to each therapeutic class. Asterisks denote BenjaminiâHochberg-adjusted q<0.05.

**Figure S8:**
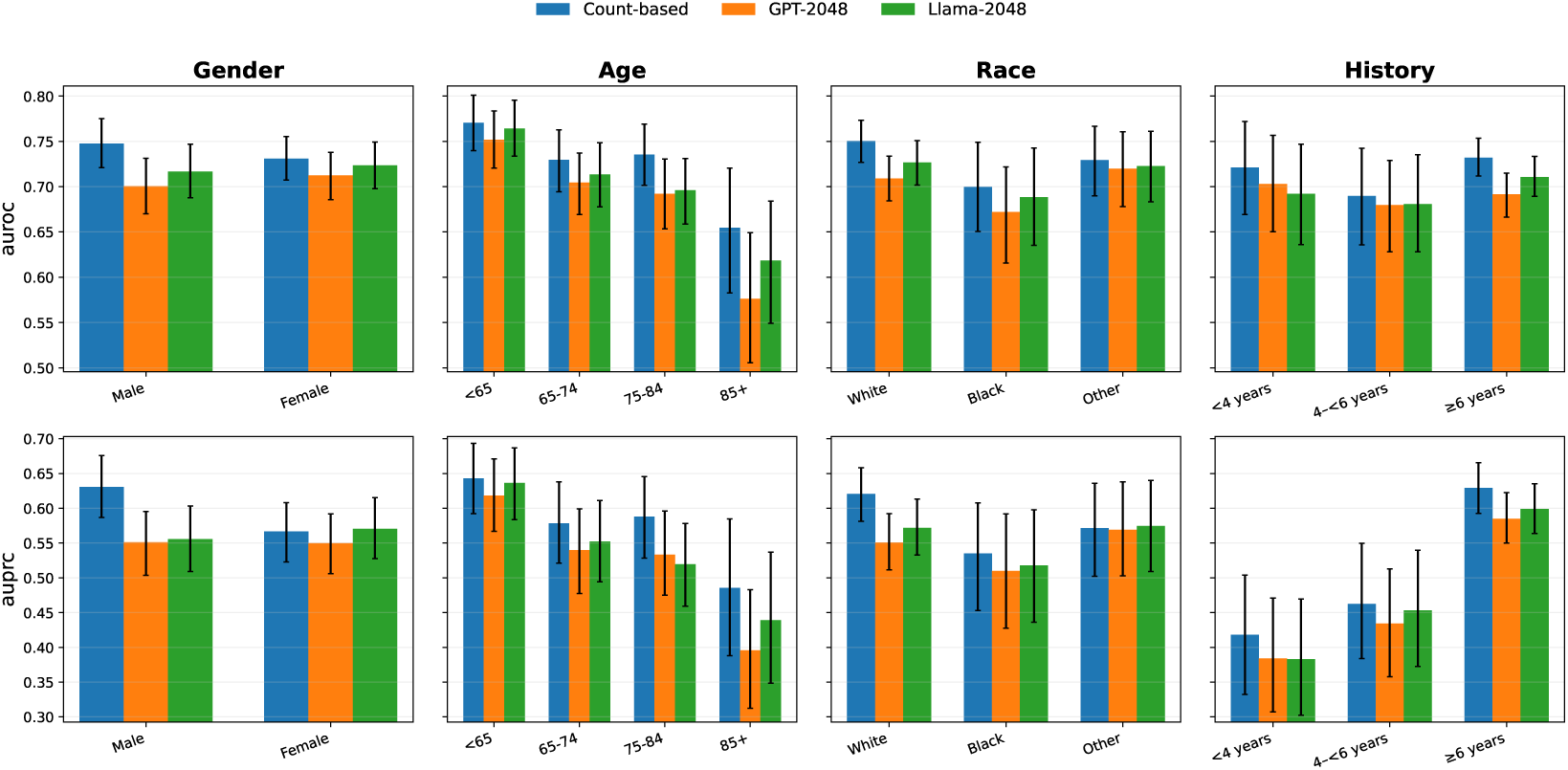
Performance across clinically relevant patient subgroups of AoU dataset. AUROC (top) and AUPRC (bottom) of the representative Count-based, GPT-2048, and LLaMA-2048 models on the held-out test set for the CD1 cohort at the 36-month prediction horizon, stratified by sex, age, race, and pre-prediction clinical history length. Error bars denote 95% bootstrap percentile intervals estimated within each subgroup.

## Footnotes

1 of mean and SD of the AUROC value

